# GM1 Gangliosidosis Type II: Results of a 10-Year Prospective Study

**DOI:** 10.1101/2024.01.04.24300778

**Authors:** Precilla D’Souza, Cristan Farmer, Jean Johnston, Sangwoo T Han, David Adams, Adam L. Hartman, Wadih Zein, Laryssa A. Huryn, Beth Solomon, Kelly King, Christopher Jordan, Jennifer Myles, Elena-Raluca Nicoli, Caroline E Rothermel, Yoliann Mojica Algarin, Reyna Huang, Rachel Quimby, Mosufa Zainab, Sarah Bowden, Anna Crowell, Ashura Buckley, Carmen Brewer, Deborah Regier, Brian Brooks, Eva Baker, Gilbert Vézina, Audrey Thurm, Cynthia J Tifft

## Abstract

**Purpose:** GM1 gangliosidosis (GM1) is an ultra-rare lysosomal storage disease caused by pathogenic variants in galactosidase beta 1 (*GLB1*; NM_000404), primarily characterized by neurodegeneration, often in children. There are no approved treatments for GM1, but clinical trials using gene therapy (NCT03952637, NCT04713475) and small molecule substrate inhibitors (NCT04221451) are ongoing. Understanding the natural history of GM1 is essential for timely diagnosis, facilitating better supportive care, and contextualizing the results of therapeutic trials.

**Methods:** Forty-one individuals with type II GM1 (n=17 late infantile and n=24 juvenile onset) participated in a single-site prospective observational study. Here, we describe the results of extensive multisystem assessment batteries, including clinical labs, neuroimaging, physiological exams, and behavioral assessments.

**Results:** Classification of 37 distinct variants in this cohort was performed according to ACMG criteria and resulted in the upgrade of six and the submission of four new variants to pathogenic or likely pathogenic. In contrast to type I infantile, children with type II disease exhibited normal or near normal hearing and did not have cherry red maculae or significant hepatosplenomegaly. Some older children with juvenile onset developed thickened aortic and/or mitral valves with regurgitation. Serial MRIs demonstrated progressive brain atrophy that were more pronounced in those with late infantile onset. MR spectroscopy showed worsening elevation of myo-inositol and deficit of *N*-acetyl aspartate that were strongly correlated with scores on the Vineland Adaptive Behavior Scale and progress more rapidly in late infantile than juvenile onset disease.

**Conclusion:** The comprehensive serial phenotyping of type II GM1 patients expands the understanding of disease progression and clarifies some common misconceptions about type II patients. Findings from this 10-year endeavor are a pivotal step toward more timely diagnosis and better supportive care for patients. The wealth of data amassed through this effort will serve as a robust comparator for ongoing and future therapeutic trials.

## Introduction

GM1 gangliosidosis (GM1) is an ultra-rare lysosomal storage disease caused by pathogenic variants in galactosidase beta 1 (GLB1; NM_000404) that result in a deficiency of lysosomal beta-galactosidase.^1,2^ Without sufficient enzyme activity, GM1 ganglioside and related glycoconjugates build to toxic levels, particularly in the central nervous system.^3–5^ The disease is primarily characterized by neurodegeneration, but systemic involvement is also present and varies across disease types and among individual patients, resulting in considerable phenotypic complexity.

The phenotype of GM1 gangliosidosis exists as a spectrum of severities that are inversely related to the residual enzyme activity and the age of symptom onset. For convenience, the phenotypic spectrum has been divided into three types.^6,7^ Type I, the infantile form (MIM #230500) is the most severe and is characterized by a developmental plateau and regression by 4–6 months of age and rapid disease progression that includes severe hypotonia, spasticity, seizures, deafness, blindness and decerebrate rigidity.^8–10^ Infantile patients with multi-organ dysfunction, including hepatosplenomegaly, cardiomyopathy, skeletal dysplasia, coarse facial features, cherry red maculae and extensive Mongolian spots^11–16^, typically have a life expectancy of 2–3 years.^17,18^ Like other lysosomal storage diseases, type I GM1 can also present as nonimmune hydrops fetalis.^10,19^ Type II (MIM #230600) is less severe and has later onset than type I, and based on the timing of observable symptom onset can be further classified into late infantile (around 1 year of age) and juvenile onset (3 – 5 years).^6,20^ Type III GM1 gangliosidosis (MIM #230650), the adult form, is the least severe. Symptoms typically emerge in the second or third decade of life.^21,22^

The available descriptions of the type II phenotype, which have recently included a wider variety of genetic backgrounds, involve developmental stagnation that evolves into ataxia, dystonia, dysarthria, and skeletal changes.^23–25^ A small cases series^26^ noted dysphagia as a leading symptom, but lower than expected incidence of visceromegaly and no facial dysmorphisms or cherry red maculae. Progressive brain atrophy and/or white matter changes on magnetic resonance imaging and progressive deficits of *N*-acetylaspartate on magnetic resonance spectroscopy have been described in type II patients.^16,27^ Clinical laboratory studies describe borderline to mildly elevated aspartate aminotransferase with normal alanine aminotransferase levels.^25^ Low bone mineral density without fractures has been documented for both subtypes, and odontoid hypoplasia has been proposed as a hallmark in late infantile disease.^28^ The largest study to date, a retrospective cohort of 41 individuals^29^, largely supported the accumulated findings. However, the available data on the natural history of type II GM1 are from samples that are either small, retrospective, or cross-sectional, which limits our understanding of the progression of disease within individual and variability therein.

Thus, we sought to add to the existing literature by conducting prospective, repeated, and comprehensive assessments of 41 participants. The aims of this report are threefold: (1) to highlight the presenting signs and symptoms of type II disease in hopes of contributing to timely diagnosis, (2) to describe the course and variability of disease facilitating better supportive care, and (3) to chronical the natural progression of disease that can serve as a control cohort for therapeutic trials.

## Methods

### Participants

Individuals with a confirmed diagnosis of GM1 gangliosidosis type II were enrolled in National Institutes of Health (NIH) clinical protocol 02-HG-0107, “Investigation of Neurodegeneration in Glycosphingolipid Storage Disorders” (NCT00029965). This study was approved by the NIH Institutional Review Board, and informed consent or assent where appropriate was obtained from all participants. Diagnosis was confirmed by review of enzyme determinations and/or biallelic pathogenic variants in *GLB1* in a CLIA-certified laboratory. Medical history and physical examinations were used to classify participants as either late infantile or juvenile onset type II GM1 gangliosidosis. A subset of the participants in this study were represented in two previous reports from this research group.^28,30^

### Clinical Assessments

This study involved in-person visits conducted at the National Institutes of Health Clinical Center (Bethesda, MD), virtual assessments conducted via phone or videoconference, and parent questionnaires completed remotely. Following an initiation visit, participants were invited to return to the Clinical Center at intervals of 1–2 years. At each study visit, participants underwent comprehensive evaluations with a multidisciplinary team, including blood, urine and/or CSF collection, abdominal ultrasound, echocardiogram (ECHO), electrocardiogram, electromyography/nerve conduction velocities, consultation with a physiatrist, audiogram and auditory brainstem response (ABR) testing, ophthalmologic evaluation, electroencephalogram, magnetic resonance imaging of the brain, speech/language assessment including a swallow study when indicated, and neurodevelopmental testing with a psychologist. Additional details about methodology are provided in the **Supplementary Methods**. Participants continued routine clinical care at their home institutions, and medication compliance and its effect on disease was not measured. General medication information is summarized in **Table 1** and in the **Supplementary Materials.** Participation in assessment was voluntary and not all patients underwent all evaluations at any given visit. Unless otherwise indicated, clinical laboratory tests were performed in the NIH Clinical Center Department of Laboratory Medicine, a CLIA-certified facility supporting the NIH Clinical Center.

**Table 1.** Participant characteristics. One participant in the Juvenile group was asymptomatic at enrollment, so age at diagnosis was less than age of symptom onset. The medication information is for the duration of the study only and does not distinguish the duration or dose (see Figure 1).

|  |  | Late Infantile Onset (n=17) | Juvenile Onset (n=24) | Full Sample (N = 41) |
| --- | --- | --- | --- | --- |
| Female, n (%) |  | 10 (59) | 14 (58) | 24 (59) |
| Race, n (%) | Asian | 1 (6) | 2 (8) | 3 (7) |
|  | Black/African American | 1 (6) | 1 (4) | 2 (5) |
|  | Multiracial | 1 (6) | 1 (4) | 2 (5) |
|  | Unknown | 0 | 4 (17) | 4 (10) |
|  | White | 14 (82) | 16 (67) | 30 (73) |
| Ethnicity, n (%) | Hispanic or Latino | 0 | 5 (21) | 5 (12) |
|  | Not Hispanic or Latino | 17 (100) | 18 (75) | 35 (85) |
|  | Unknown | 0 | 1 (4) | 1 (2) |
| Country of residence, n (%) | Germany | 0 | 1 (4) | 1 (2) |
|  | Poland | 0 | 1 (4) | 1 (2) |
|  | United States | 13 (76) | 22 (92) | 35 (85) |
|  | Russia | 1 (6) | 0 | 1 (2) |
|  | Sweden | 3 (18) | 0 | 3 (7) |
| Proband, n (%) |  | 12 (71) | 18 (75) | 30 (73) |
| Age at consent (years) | Median [IQR] | 4.98 [3.52, 6.09] | 12.02 [7.18, 15.01] | 7.03 [4.98, 12.27] |
|  | Range | 2.61 - 8.72 | 1.41 - 23.78 | 1.41 - 23.78 |
| Age at symptom onset (years) | Median [IQR] | 1.5 [1.08, 1.5] | 3 [2.5, 4] | 2.5 [1.5, 3.5] |
|  | Range | 1 - 2 | 1.5 - 6 | 1 - 6 |
| Parent-reported symptoms at onset<sup>a</sup>, n (%) | Motor | 15 (88) | 16 (67) | 31 (76) |
|  | Speech | 14 (82) | 21 (88) | 35 (85) |
| Age at diagnosis (years) | Median [IQR] | 3 [2.33, 3.75] | 9.5 [6.69, 11.25] | 6 [3, 10] |
|  | Range | 1.25 - 6 | 1 - 19 | 1 - 19 |
| Gastronomy tube, n (%) |  | 9 (53%) | 8 (33%) | 17 (41%) |
| Age at G-tube, (years) |  | 5 [5, 6] | 17.5 [15.5, 21] | 9 [5, 17] |
| Any miglustat during study, n (%) |  | 4 (24) | 10 (42) | 14 (34) |
| Any tanganil during study, n (%) |  | 2 (12) | 2 (8) | 4 (10) |
| Any seizure meds during study, n (%) |  | 11 (65) | 7 (29) | 18 (44) |

<sup>a</sup>Two juvenile participants are missing this information, so juvenile N = 22. Categories are not mutually exclusive.

### Analytic Approach

The goal of this study is to describe the course and variability of the signs and symptoms of type II GM1. Given the differential course of illness, in most cases results are reported separately for the late infantile and juvenile onset groups. The available longitudinal data are summarized graphically to illustrate the course of illness. However, the sample size could not support the appropriate inferential models (e.g., generalized linear models). For this reason, we focus on the descriptive analysis (frequencies for categorical data, median and interquartile ranges for ordinal or skewed data, and means and standard deviations for normally distributed data). To additionally support interpretation about the course of the disease using cross-sectional information, for quantitative outcomes we provide Spearman correlations with chronological age (ρ_age_), or for categorical outcomes the distribution of age within each level.

## Results

### Participant Characterization

In total, 41 individuals with GM1 gangliosidosis type II were evaluated, including eight sibling pairs or trios (**Table 1**). Participants were classified as late infantile (10 female, 7 male) or juvenile (14 female, 10 male) onset. **Figure 1** summarizes the timing of key events.

**Figure 1.**
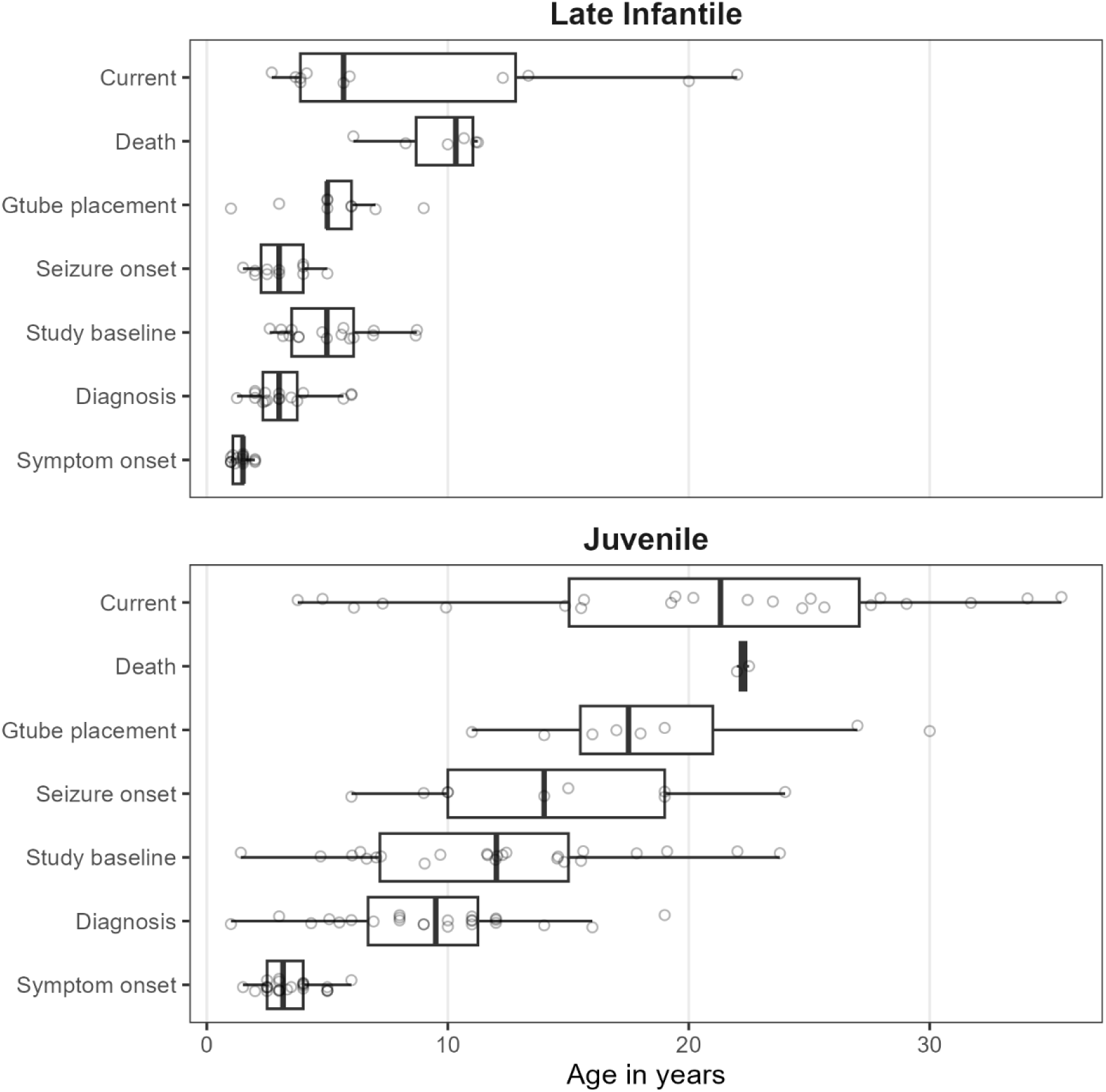
Summary of participant data. The timing of disease milestones and study participation are plotted. “Symptom onset” refers to the parent-reported age of first symptoms, which were either motor or speech-related. “Current age” refers to the age of last contact for participants who were alive. For most participants, this was August 2023, but for those who could not be recontacted it was the last visit date and for those enrolled in the gene therapy trial, it was the enrollment date of that study.

#### Genotype

Likely causative variants were identified in *GLB1* in all individuals enrolled in the study and classified according to ACMG guidelines^31^ and modified criteria.^32–35^ Most participants were compound heterozygous for pathogenic (PATH) or likely pathogenic (LPATH) variants in GLB1 (late infantile: 12, 71%; juvenile: 22, 96% of 23 who were genotyped). In total, 37 variants were identified and classified; 30 were classified as PATH and seven as LPATH (see **Supplementary Tables S1**). In ClinVar, six variants were upgraded from VUS to LPATH, and four previously unreported variants were submitted, all classified as PATH or LPATH.

#### Symptom onset and diagnosis

Retrospective report of developmental milestones was available for 16 (94%) late infantile and 24 (100%) juvenile onset participants. Those with late infantile onset usually exhibited delay or non-acquisition of major milestones (sitting, walking, first words), subsequently losing whatever skills had been acquired. However, the juvenile onset group usually attained these milestones on time relative to population norms and while loss was less common than for late infantile onset, it was not rare (**Supplementary Figure S1**). Most participants in both groups experienced both motor and speech-related initial presenting symptoms (**Table 1**). The median [IQR] time from symptom onset to diagnosis in probands (i.e., excluding siblings) was 1.53[0.96, 2.13] years for late infantile probands (n=12) and 5.5[3.42,8.75] years for the juvenile probands (n=18).

### Physical Metrics

#### Ophthalmology

Seventeen (100%) of the late infantile participants had at least one ophthalmological evaluation; the most recent result for each participant is reviewed here. Among the late infantile cohort, only six were able to complete formal acuity testing with a Snellen equivalent, with an average LogMAR acuity of 1.20 (equivalent to 20/314) (the remaining were recorded as no or questionable blink to light, n=3; blink to light, n=4; occasional fix/follow, n=1; fix/follow, n=1; and central/steady/maintained or central/steady/unmaintained, n=2). Ptosis was rare and mild-to-moderate (n=1, 6%), but strabismus and nystagmus were common (n=17, 100% and n=9, 53%, respectively). Mild (n=8, 47%) and moderate (n=1, 6%) corneal clouding (hazing) was observed. On retinal exam no patients had a cherry-red maculae, but optic nerve pallor/atrophy was noted in four (24%) late infantile patients. The mean spherical equivalent of the refractive error was +2.62±2.53 Diopters (n=17). Cortical/neurological vision impairment was notable in 12 (71%).

Twenty-one (88%) juvenile participants had at least one ophthalmological evaluation. Their visual acuity ranged between 20/20 and 20/200; the average LogMAR acuity for the better seeing eye was 0.34 (equivalent to 20/44) (n=19 with available data). No nystagmus was noted in the juvenile cohort, but mild-to-moderate ptosis (n=4, 19%) and strabismus (n=5, 24%) were observed. Corneal clouding/hazing (mild, n=9; moderate, n=3) was common. Retinal exam was generally normal, with occasional granular retinal periphery which may be normal variant, and optic nerves were mostly normal (only the oldest patient exhibited mild degrees of pallor). No cherry red maculae were observed. Refractive error was mostly within normal range for age (mean spherical equivalent of the refractive error, +1.10 ± 2.64 Diopters). Finally, no cortical/neurological visual impairment was observed.

#### Audiology

All participants received at least one audiological evaluation and their most recent and most complete assessment was used for cross-sectional summary (**Supplementary Table S2**). Peripheral hearing sensitivity was within normal limits for most participants (late infantile: n=15, 88%; juvenile: n=22, 92%), and the available longitudinal data (late infantile: n=6, 35%; juvenile: n=16, 67%) indicated that it was stable. There was no evidence of gross middle ear dysfunction in any participant. Among those who received neurodiagnostic ABR testing, the results were within normal limits for eight of 15 (53%) late infantile and 14 of 22 (64%) juvenile participants. Abnormalities included small V/I amplitude, poor neural synchrony, and/or retrocochlear auditory dysfunction. No gross changes were noted in the available longitudinal data, indicating stability.

#### Video fluoroscopic assessments of swallowing (VFSS)

For 14 (82%) of the late infantile group and 23 (96%) of the juvenile group, VFSS was prompted by clinical interview and assessment (age in years at assessment median[IQR]: late infantile, 5 [3.5, 5.9]; juvenile, 11.8 [7.6, 14.7]). In the juvenile group, VFSS scores (see **Supplementary Table S6**) indicated little risk of aspiration (median[IQR]: 5 [4, 5], ρ_age_ =-0.28) and safe and efficient swallowing (median[IQR]: 4 [3.5, 4.5], ρ_age_ =-0.53). Dietary restrictions were generally absent or minimal (median[IQR]: 4 [4, 5], ρ_age_ =-0.68), and only three participants (aged 15 – 24 years) had maximal or moderate restriction. Aspiration (median[IQR]: 4 [0.25, 5], ρ_age_ =-0.35) and swallowing (median[IQR]: 3 [0.25, 4.75], ρ_age_ =-0.36) scores were slightly more severe and variable in the late infantile group. For the majority of late infantile participants (n=9 of 14), dietary restrictions were minimal or none, but the remaining five participants had maximal restriction (median[IQR]: 4 [2, 5], ρage=-0.44). The negative correlations with age reflected more severe ratings in older participants than in younger participants, but the limited within-participant longitudinal data indicated stability over the observation period.

#### Cardiology

ECHO was performed for nine (53%) late infantile participants, revealing only one with mitral valve prolapse and associated mitral regurgitation. No evidence of aortic valve leaflet thickening or dilated or hypertrophic cardiomyopathy was observed. One participant did have borderline aortic root dilation. Among 13 juvenile participants receiving ECHO, three (23%) had aortic valve leaflet thickening with associated aortic valve regurgitation ranging from trace to moderate in severity (**Figure 2**). One of these affected individuals also had associated trivial aortic stenosis. Five additional participants had aortic and/or mitral valve leaflets that were qualitatively thickened for age without any associated valvular regurgitation or stenosis but did not reach a threshold to be commented upon by the clinical cardiology teams. Biventricular function and chamber dimensions remained within acceptable limits for age in all cases. No clinically significant EKG abnormalities were observed among the participants who were evaluated (late infantile: n=9, 53%; juvenile: n=15, 63%).

**Figure 2.**
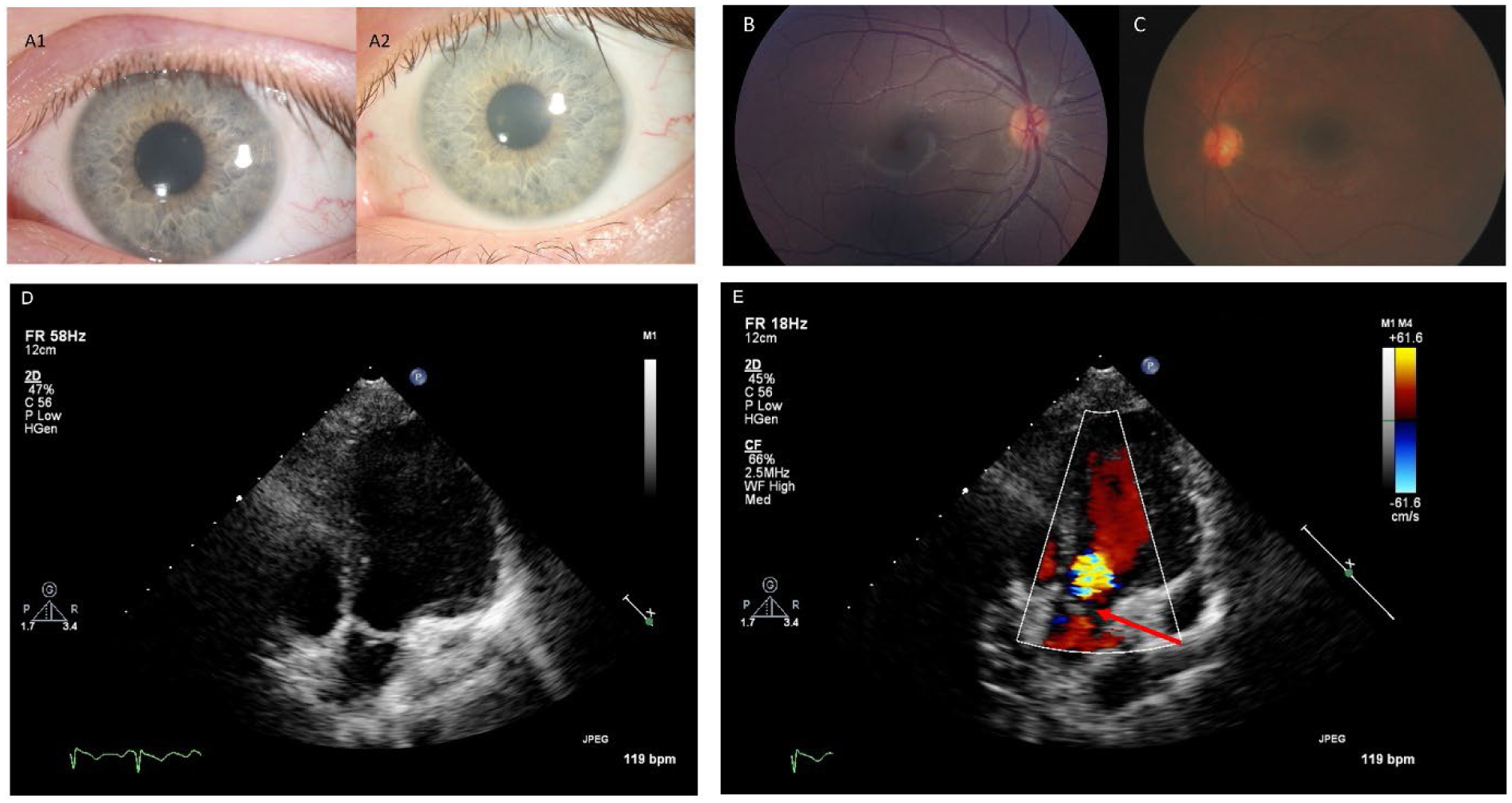
Selected ophthalmology and cardiology results. Panels A1 and A2: Corneal slit-lamp photographs from the same Juvenile GM1 patient showing slow progression of corneal clouding over a five-year period. Panel B: Left eye Topcon color fundus photograph from a Juvenile GM1 patient showing an essentially normal fundus with no evidence of macular or optic nerve head pathology. Panel C: Right eye RetCam fundus photography of a late-infantile GM1 patient showing normal macula and optic nerve with minimal retinal vascular tortuosity. Panels D and E: Two dimensional and color transthoracic echocardiographic imaging of the left ventricular outflow tract from a single Juvenile GM1 patient. The red arrow points to aortic valve leaflet thickening with associated aortic regurgitation. These images are representative of the abnormal thickening of the aortic valve leaflets with associated aortic regurgitation observed in three unrelated Juvenile GM1 patients (23% of those who received ECHO; an additional five participants had aortic and/or mitral valve leaflets that were qualitatively thickened for age but without regurgitation or stenosis).

#### Abdominal Ultrasound

At least one abdominal ultrasound was performed in nine late infantile (53%; age median[IQR] = 3.5[3.8, 6.1]) and 17 juvenile participants (71%; age median[IQR] = 7.2[12.3, 19.9]). The majority of assessments were normal; at the first available timepoint, mild enlargement was observed for two (22%) late infantile participants and two (13%) juvenile participants. All four participants with radiologically abnormal findings had abnormal AST levels, one had abnormal ALT, and none had abnormal GGT. The two late infantile participants with enlargement had no follow-up, but each of the juvenile participants had a 1-year follow-up showing hepatomegaly for one and a return to normal for the other. None of the participants with hepatomegaly had splenomegaly, however, minimal-to-mild splenomegaly was observed at the first assessment for two other late infantile participants (22%) and one other juvenile participant (6%). No follow-up imaging was available for these three individuals. An additional juvenile participant had developed mild splenomegaly 1 year after the first evaluation.

#### Mobility

At least one mobility assessment was available for all participants (**Supplementary Figure S2**). In the late infantile group, baseline floor mobility scores were variable, with a median score of “sits without support” (median[IQR] = 3 [0, 5]), and older participants had worse scores than younger participants (ρ_age_ = −.47). In the juvenile group, baseline upright mobility scores were less severe, with a median score of “independent ambulation, may be unsteady” (median[IQR] = 4 [1.75, 4]). Scores in the juvenile group were also strongly correlated with age (ρ_age_ = −.74). The longitudinal data indicated worsening over time within person for both onset groups (**Supplementary Figure S2**). To support the validity of the mobility assessment, we compared the scores with the Vineland-3 Gross Motor GSVs for 21 participants with contemporaneous evaluations, finding strong positive Spearman correlations (floor, ρ=0.87; upright, ρ=0.75).

### Laboratory Studies

The β-gal enzyme activity in available serum for late infantile (n=11, 65%) and juvenile (n=21, 88%) patients ranged from 0–5% (median[IQR] = 0.028[0.014, 0.04]) of the pediatric control sample (**Supplementary Figure S3**). Enzyme activity in available CSF for late infantile (n=5, 29%) and juvenile (n=13, 54%) patients ranged from 2–8% (median[IQR] = 0.05[0.026, 0.07]) of the same pediatric controls. Serum and CSF were not meaningfully correlated with one another, though this was likely due to the extremely limited ranges (**Supplementary Figure S3**).

Blood samples from all participants were evaluated for complete blood count with differential, complete metabolic panel (hepatic panel, creatinine kinase, lipid panel), iron panel, thyroid panel, vitamin D, prothrombin time/partial thromboplastin time, and lactate dehydrogenase. Within the cross-sectional data (first available visit), all parameters were in the normal pediatric ranges except for several liver enzymes: aspartate aminotransferase (AST) was elevated for 13 (76%) late infantile and seven (29%) juvenile onset patients, alanine aminotransferase (ALT) was elevated for three (18%) late infantile two (8%) juvenile onset patients, and gamma-glutamyl transferase (GGT) was elevated for six (35%) late infantile and six (25%) juvenile onset patients (**Supplementary Table S3**). The longitudinal data suggested stability in these parameters, indicating that these parameters did not change systematically within individuals over the observation period (**Supplementary Figure S4**).

### Electrodiagnostic and Neuroimaging

#### Electroencephalography (EEG)

Twelve (71%) late infantile participants received at least one outpatient ambulatory EEG; the first available EEG is summarized here (age at first assessment, median 5.3[3.4, 6.9] years). Most (n=10, 83%) of these EEGs were abnormal: background abnormalities (including low overall voltage, slow posterior dominant rhythm or abnormal background slowing) were noted in nine (90%), focal or persistently rhythmical brief slowing was noted in four (40%) (delta-range predominant in two, theta-range predominant in two), and four (40%) had epileptiform activity (typically sharp waves over temporal and sometimes frontal head regions). One of the participants with a normal EEG at the first visit had possible progression with the lack of a normal posterior basic rhythm in an overnight EEG 1 year later; the other participant did not have any follow-up EEGs. Four of the participants with an abnormal first EEG had repeated ambulatory or overnight assessments, with consistently abnormal results.

The 18 (75%) juvenile participants with at least one EEG had a median age of 15.2[11.7, 17.9] years at the earliest available assessment. All 12 (67%) abnormal EEGs had background abnormalities, five (42%) had focal or persistently rhythmical bursts of brief slowing (theta-range in four patients, and alpha-range in one patient), and one (6%) had epileptiform activity (typically sharp waves over temporal and sometimes frontal head regions). Among the six participants with normal EEGs at the earliest assessment, two had an abnormal EEG (background abnormalities, similar to those noted in the late infantile onset group) at a later timepoint, one had two more normal EEGs, and three had no other EEGs. Among the 12 participants with abnormal EEGs at the earliest assessment, six had no follow-up ambulatory or overnight EEG, four had two or more follow-up ambulatory or overnight EEGs that were consistently abnormal, and two had both normal and abnormal follow-up EEGs.

#### Structural magnetic resonance imaging (MRI) of the brain

Out of 14 (82%) of the late infantile cases with at least one MRI, 11 (79%) presented with or developed atrophy of the cerebellum. Two of these cases were described as moderate, and nine as mild atrophy. Eleven (79%) late infantile participants presented with or developed cerebral cortical atrophy, including four severe and three moderate. A mid sagittal T1-weighted image was not available for one participant, so the size of the brainstem and corpus callosum could not be evaluated in this case. Of the remaining 13 cases, five (38%) presented with or developed brainstem atrophy, including one severe, and nine (69%) with atrophy of the corpus callosum, including five severe and two moderate. Cerebral white matter could be assessed in only eight cases (other cases lacked the pulse sequences necessary for proper evaluation of myelination) all showed abnormalities of myelination, half (n=4) of which were severe. Using the latest available observation for each person, the qualitative atrophy scores in each of these regions was correlated with age, such that older participants had worse ratings (cerebral cortex, ρ_age_ = 0.79; corpus callosum, ρ_age_ = 0.72; see also **Figure 3**). Longitudinal data suggested that most participants worsened over time (**Figure 3**).

**Figure 3.**
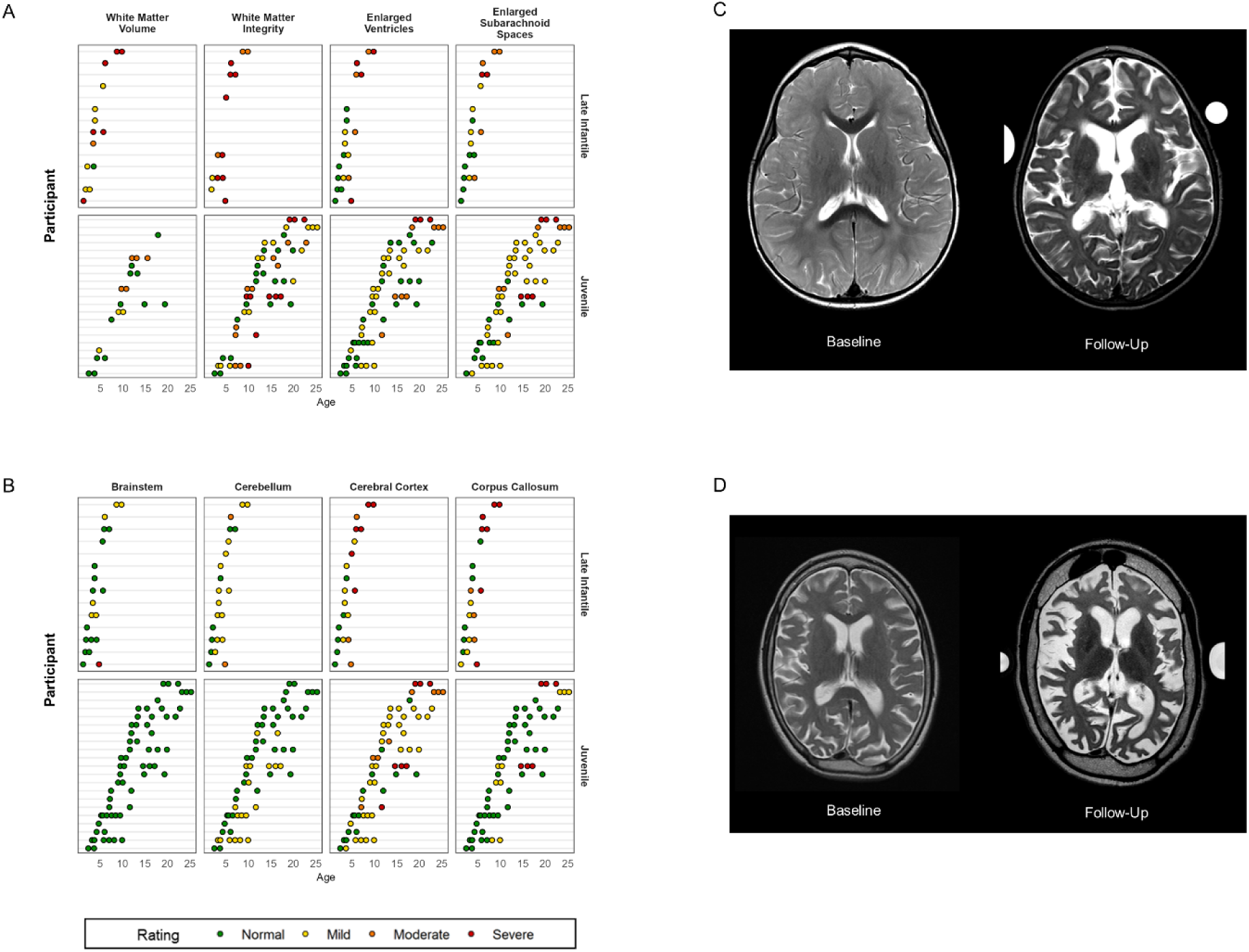
MRI qualitative ratings for all participants. Panels A and B: Qualitative MRI ratings are shown per MRI per person (Y-axis) by age in years (X-axis). Late infantile onset data are shown in the top rows and juvenile onset data in the bottom row. Cerebral white matter could only be assessed in n=8 participants with late infantile onset, as the others lacked the pulse sequences necessary for proper evaluation. Panel C: Serial MRIs from a single patient with late infantile onset at baseline and a follow-up 2.5 years later (ages redacted per MedArXiv requirements). Panel D: Serial MRs from a single patient with juvenile onset baseline and a follow-up 8 years later (ages redacted per MedArXiv requirements).

Twenty-one (88%) of the juvenile participants had at least one MRI. Six (29%) presented with or developed mild atrophy of the cerebellum; 17 (81%) with cerebral cortical atrophy, including three severe and three moderate; and five (24%) with atrophy of the corpus callosum, including two severe. Out of 19 cases for which white matter could be evaluated, 13 (68%) presented with or developed white matter injury, including four severe and five moderate. The brainstem appeared normal in all cases. No consistent cross-sectional trends between age and atrophy were observed using the most recent MRI, and the available longitudinal data indicated general stability for most participants during the observation period (**Figure 3**).

#### Magnetic resonance spectroscopy (MRS) of the brain

At least one visit with MRS was available for 13 (76%; n=2 with two visits) of the late infantile group and 21 (88%; n=14 with two, three, or four visits) of the juvenile group. For both groups, the average concentration of creatine, choline, and glutamine+glutamate+gamma-aminobutyric acid (glx) measured in the left centrum semi-ovale were similar to age-normative expectations, but both myo-inositol (elevated) and *N*-acetylaspartate+*N*-acetylaspartyl glutamate (NAA; decreased) were abnormal (**Figure 4 and Supplementary Table S4**). Within the cross-sectional data, older age was associated with increased deviation from normative values for both myo-inositol (late infantile ρ=0.28; juvenile ρ=0.68) and NAA (late infantile ρ=-0.59; juvenile ρ=-0.74) (see **Supplementary Table S4a**). The juvenile cohort had sufficient data to attempt formal longitudinal modeling (see **Supplementary Methods**) (**Supplementary Figure S5**). Consistent with the cross-sectional analysis, older participants tended to have greater excess of myo-inositol (t=2.63, p=0.016) and greater deficit of NAA (t=-3.56, p=0.002) than younger participants (see **Supplementary Table S4b**). However, a trend towards detectable within-person change relative to normative expectations during the observation period was observed only for myo-inositol (t=2.01, p=0.068) and not for NAA or other metabolites.

**Figure 4.**
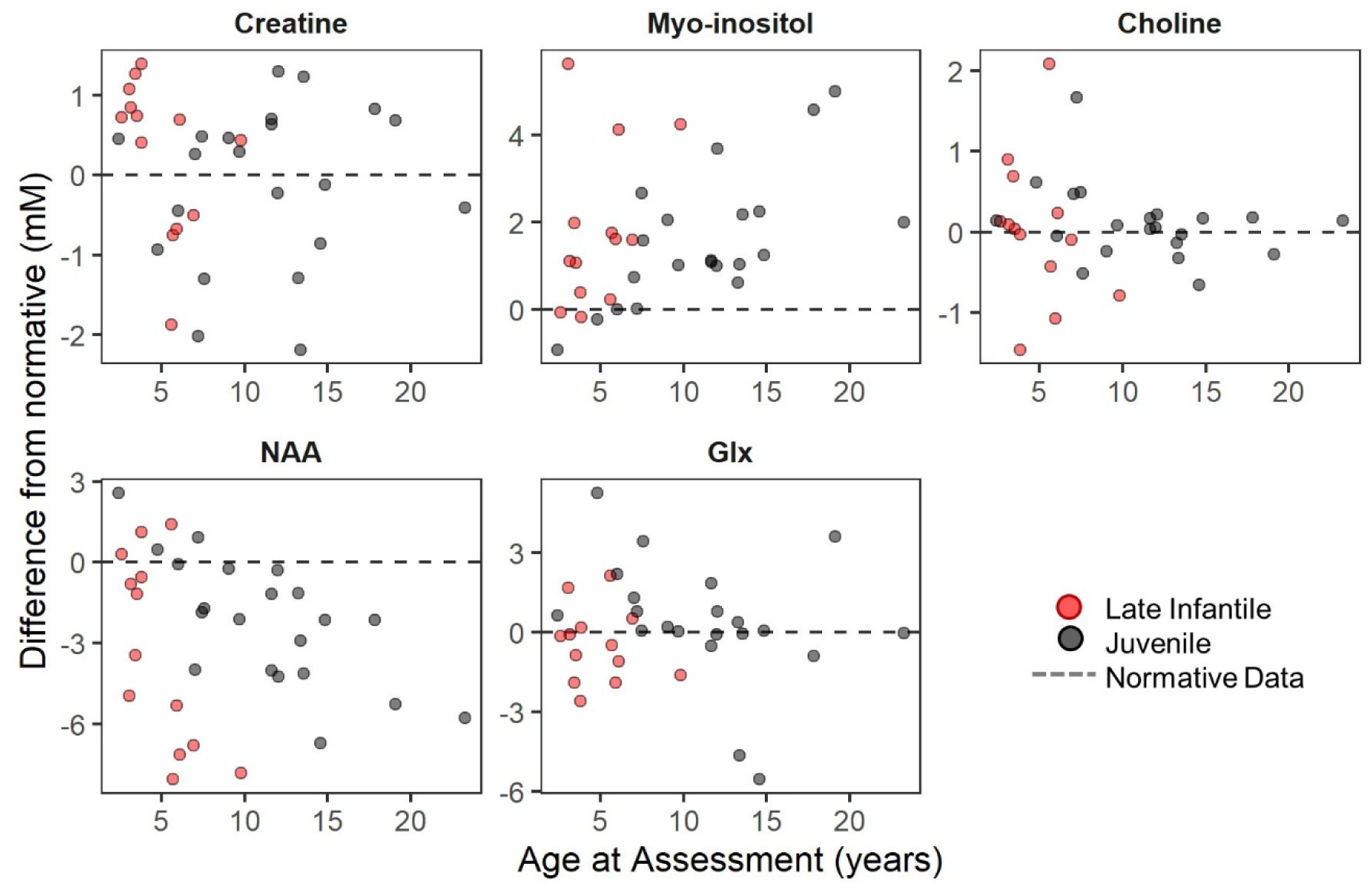
Age progression of metabolite concentration (mM) in LCSO relative to normative expectations. NAA: *N*-acetyl aspartic acid. Glx: Glu+Gln+GABA. Red = late infantile onset; black = juvenile onset. Earliest available observation per person (Late Infantile, n = 13; Juvenile, n = 21) is plotted. Points show difference in concentration from age expectation, which is illustrated by the dotted line at zero.

### Neurodevelopmental Assessments

#### Speech evaluation

All participants in both groups participated in at least one speech evaluation (see **Supplementary Table S6**); the earliest available assessment is reported here (median[IQR] for age at assessment: late infantile, 5 [3.5, 5.9]; juvenile, 11.8 [7.6, 14.7]). Most (n=13, 76%) of the late infantile onset group had the most severe score of zero for both speech and language ratings (median[IQR] scores: speech, 0 [0, 0]; language, 0 [0, 0]). Three late infantile participants had longitudinal data, and their scores were at the floor for all visits. The juvenile onset group had a severe speech presentation (median[IQR]= 2 [1, 3]) and was deficient in most areas of language (median[IQR]= 1 [1, 1]); both of these scores were more severe in older participants than in younger participants (correlation with age: Speech ρ = −0.60; Language ρ = −0.53). The limited juvenile within-subject longitudinal data indicated stability (**Supplementary Figure S6**).

#### Adaptive behavior

Most late infantile (n=14, 82%) and juvenile (n=23, 96%) participants had at least one administration of the Vineland Adaptive Behavior Scales (see **Supplementary Figure S7** for longitudinal data). The cross-sectional dataset was created using the participant’s first Vineland-3 (Late infantile, n=8; Juvenile, n=14), or their first Vineland-II (Late infantile, n=6; Juvenile, n=9) if the third edition was not given. Most participants in both groups (Late infantile: n=12 of 14; Juvenile: n=19 of 23) had Adaptive Behavior Composite (ABC) standard scores below the 2.5th percentile. The median ABC scores were in the moderately impaired range for both groups, and floor effects were common for the juvenile onset group, but not the late Infantile group (**Table 2**; see also **Supplementary Table S5**). Strong negative correlations were observed between ABC scores and chronological age in both groups. **Figure 5** illustrates that floor effects in V-scale scores dramatically obscured the observed between- and within-participant variability in performance, and that the growth scale values (GSVs) were sensitive to that variability, making the latter more useful as outcomes in future trials.

**Figure 5.**
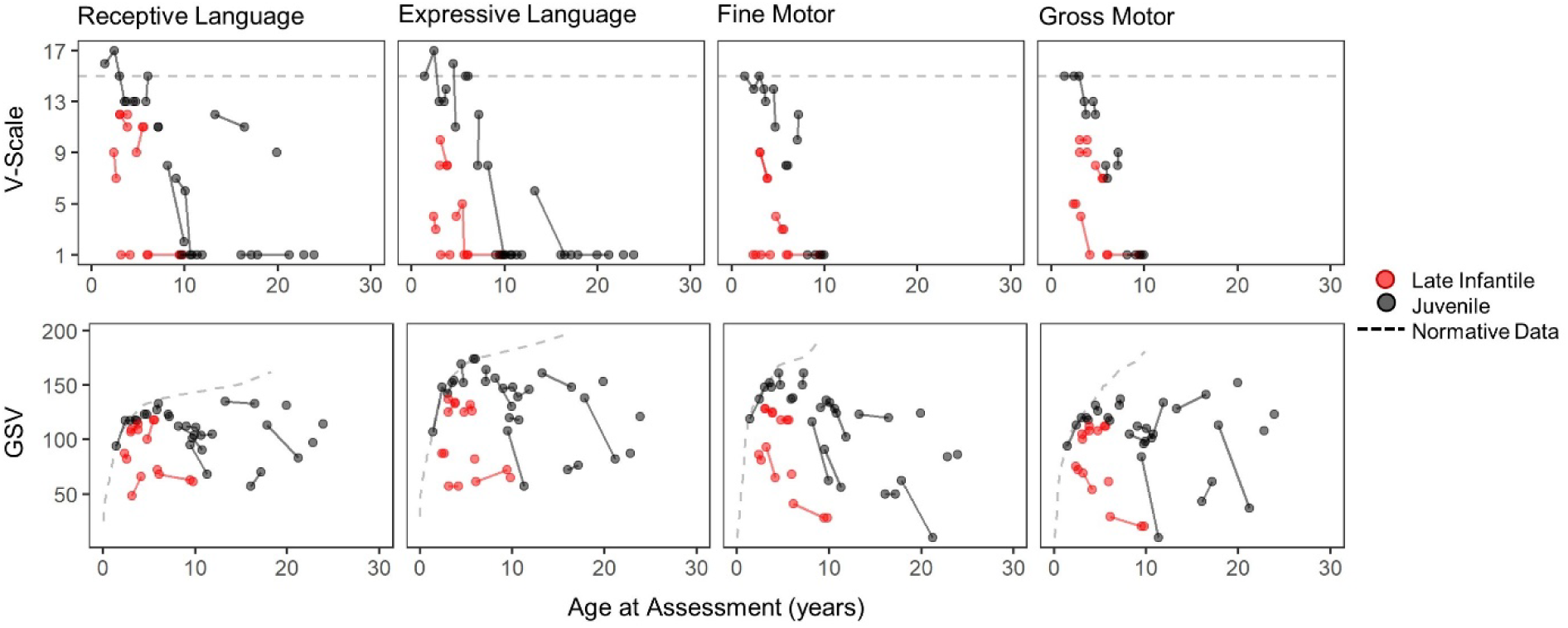
Selected Vineland-3 subdomain scores, expressed as V-scale (top row) and growth scale value (GSV; bottom row). Red = late infantile onset (n = 8); black = juvenile onset (n = 14). Solid lines connect observations from the same person. V-scale scores have a population mean of 15 (dotted line) and SD of 3. The population-level distribution of GSV is not defined, but the median value per age equivalent in the normative sample is plotted (dotted line). Gross and Fine Motor subdomain V-scale scores are available only through age 9 years.

**Table 2.**
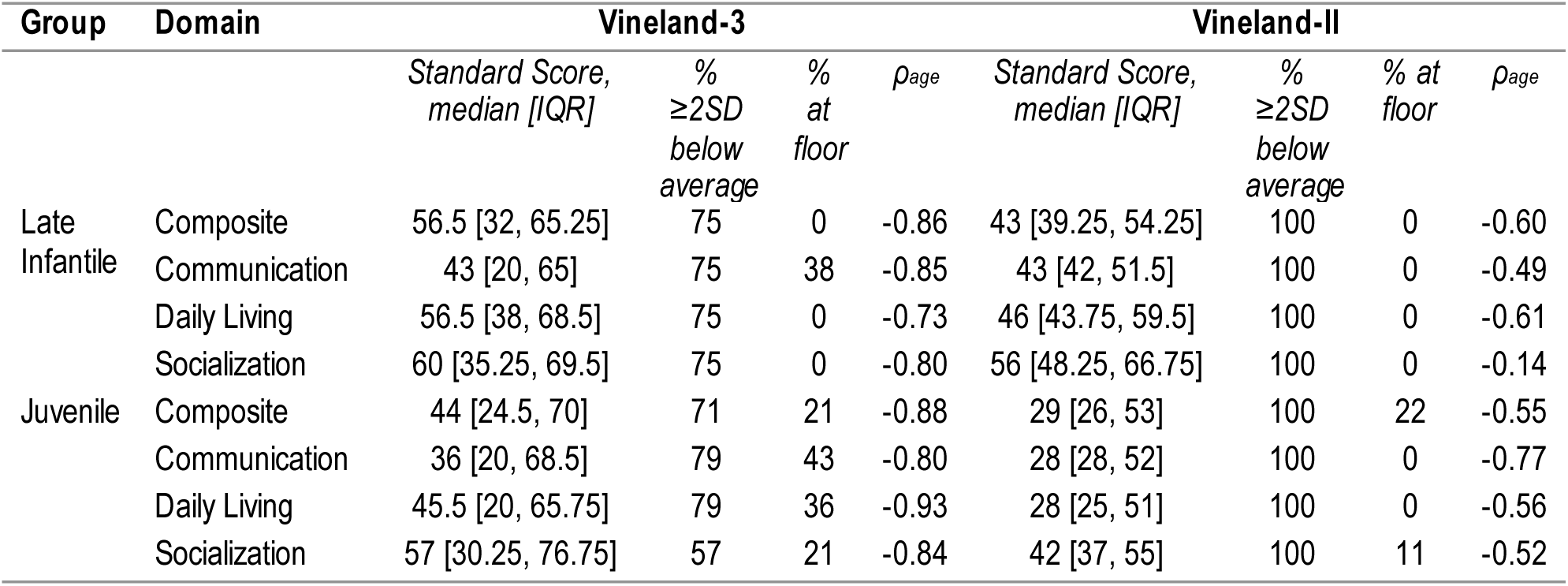
Vineland Adaptive Behavior Standard Scores at the First Visit. The first Vineland-II and/or first Vineland-3 assessment available for each person is summarized in this table. The Spearman correlation between the standard score and age is shown in the *ρ_age_* column. Sample sizes and ages were: Late Infantile/Vineland-3: n = 8 (median age: 3.97 [3.06, 5.98]), Late Infantile/Vineland-II: n = 6 (median age: 4.09 [3.46, 4.91]); Juvenile/Vineland-3: n = 14 (median age: 9.59 [7.37, 17.39]), Juvenile/Vineland-II: n = 9 (median age: 17.29 [11.99, 19.38]). Motor excluded as standard scores are not available for all participants. Standard scores have a population mean of 100 and SD of 15 (floor = 20). IQR = interquartile range (25th – 75th percentile).

We explored the relationship between MRS, a potential severity biomarker, and adaptive behavior scores. Given that the MRS concentrations are expressed as deviance from age-expected values, we compared those to the age-norm-referenced ABC standard scores. Using the cross-sectional data (first available Vineland-3, n=8 late infantile and n=13 juvenile, otherwise first available Vineland-II, n=4 late infantile and n=6 juvenile) we calculated Spearman correlations between ABC and MRS (**Figure 6**). Myo-inositol elevations (ρ_VABS3_=-0.90, ρ_VABSII_=-0.61) and NAA deficits (ρ_VABS3_=0.64, ρ_VABSII_=0.64) were strongly correlated with more impaired adaptive behavior.

**Figure 6.**
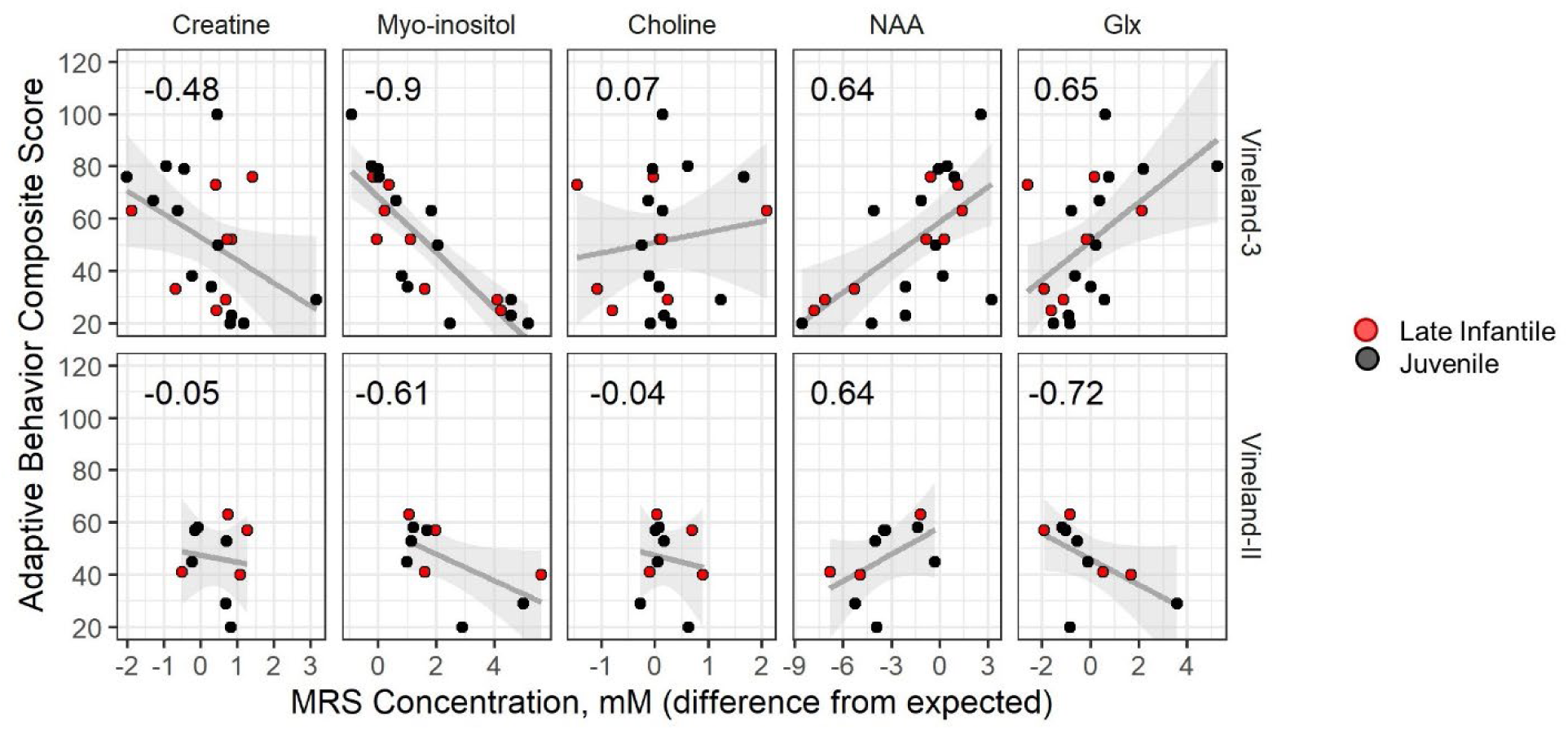
Correlation between LCSO MRS concentration (mM, deviation from expected) and Vineland scores. Cross-sectional data are plotted; the earliest available Vineland-3 was selected for each participant (top row) and if the participant had no Vineland-3 then the earliest available Vineland-II was used (bottom row). Each panel illustrates the Spearman correlation (value inset) for the relationship between Vineland Adaptive Behavior Composite (ABC) and the MRS concentrations (mM) in LCSO, each expressed as difference from age expectations (columns). Correlations are shown separately for the second and third editions of the Vineland as the scores are not combinable. Points are colored by onset group, black is late infantile and red is juvenile. Gray lines and shaded area represent the linear regression line and 95% confidence interval.

## DISCUSSION

The comprehensive serial phenotyping of type II GM1 patients in this 10-year prospective study expands our understanding of the disease spectrum. While the classification of disease into late infantile and juvenile onset parses some heterogeneity in presentation and progression, considerable variability remained even within the subtypes. Although small sample size precludes a genotype-phenotype investigation, it seems reasonable to assume that this heterogeneity is related to the high degree of genetic variability and compound heterozygosity among the 165 pathogenic and 112 likely pathogenic variants in *GLB1* described in this disease.^36^ Using ACMG criteria we classified each of the 37 variants in our cohort; all of these were classified as pathogenic or likely pathogenic. The classification is based on a deeply phenotyped cohort and low enzyme activity in serum and CSF compared to an age-matched pediatric control group. We anticipate that this classification will aid clinical laboratories in variant interpretation when the phenotype or enzyme testing results are unavailable.

As is often documented for other ultra rare genetic conditions associated with neurodevelopmental disability^37^, delay, absence, or loss of major milestones such as walking and talking were typically the first observable symptoms of type II disease. These symptoms are perhaps more subtle, and certainly less specific, than the various physical manifestations described as cardinal symptoms of type I disease.^11–16^ This may explain the long delay between symptom onset and diagnosis in this study – 1.5 years for late infantile onset and 5.5 years for juvenile onset probands. Loss of major milestones, which occurs very rarely among healthy children, was common in both late infantile and juvenile onset groups. Further, the parent-reported and direct assessments in this study converged upon a clinical picture of ongoing deterioration in whatever gross motor and language abilities had been attained for both onset groups, though the progression was more rapid among late infantile patients. Thus, loss or deterioration in motor or speech domains is a particularly salient indicator for genetic testing, even if milestone attainment had previously been normal.

The combination of retrospective reports and prospective evaluations in this study yielded valuable insights into two domains of clinical concern in GM1, feeding and seizures. Consistent with other reports^29^, gastronomy tubes were common among participants with late infantile onset, most of which were placed in early childhood. Only about 1/3 of juvenile onset participants received a gastronomy tube, but the placement age was usually late in the second decade of life. The prospective formal assessment of swallowing behaviors revealed that older participants tended to have worse swallowing abilities. Because feeding is a clinically salient symptom domain, these findings support the use of gastronomy tube placement and/or more precise quantification with swallowing evaluations as outcome measures in future study.

Seizures are a hallmark of GM1, and it is known that most patients will develop clinical or electrographic seizures over the course of their disease. Indeed, nearly all late infantile onset participants had a history of seizures prior to enrollment, with several experiencing seizures prior to the GM1 diagnosis, and all participants who developed seizures did so by age 5 years. Seizures were less common, but not rare, for the juvenile onset group. However, the onset age was extremely variable, ranging from 6 to 24 years. While the retrospective report of clinical diagnosis of seizure is important, it remains possible that atypical presentation may prevent the timely diagnosis of seizures in individuals with GM1. Because the deep phenotyping of this study included prospective EEG evaluation, we were able to document that 83% of late infantile and 67% of juvenile EEGs were abnormal either due to background abnormalities, focal slowing, or epileptiform activity.

The prospective design of this study included evaluations of other areas thought to be impacted by GM1. Upon systematic ophthalmologic examination, no participants in this cohort exhibited cherry red maculae, though the extent of other ophthalmologic abnormalities appeared to be related to the severity of disease. Similarly, while hearing impairment has been described as a feature of Type I GM1^2^, the majority of Type II participants in this study had normal hearing sensitivity that was stable over time, with no middle ear dysfunction and normal ABRs. Thus, the absence of cherry red maculae or hearing impairment should not distract the diagnostic odyssey from the GM1 diagnosis.

Although cardiomyopathy is common in other LSDs such as Fabry disease and Pompe disease^38,39^ the results of the current study suggest that it is not a characteristic feature of type II GM1 gangliosidosis. About half of this cohort underwent cardiac ultrasound exam, yielding essentially normal results. However, three of the oldest juvenile onset patients had aortic valve leaflet thickening with regurgitation suggesting that for older type II juvenile onset patients, periodic assessment of cardiac function may be indicated.

Hepatosplenomegaly is a hallmark of some LSDs and in a recent retrospective natural history report was identified in the majority of early onset GM1 patients.^11,18,29^ Interestingly, it was not a major feature in our type II GM1 patients, as only mild or borderline enlargement observed in only a minority of patients upon systematic direct assessment. Still, liver enzymes GGT, ALT, and AST indicated some (stable) elevations in many patients. AST was elevated in all the participants with liver enlargement, but it was also elevated in most other late infantile onset patients and about 30% of the juvenile patients. Elevations in AST most often reflect liver dysfunction particularly in association with other liver enzymes ALT and GGT. However, AST evaluations can also reflect other sources including cardiac, muscular, hematologic, renal, and endocrine systems ^40,41^ and acute encephalopathy and seizures.^42^ The elevation of additional liver enzymes such as elevated GGT in 35% of the late infantile and 25% of the juvenile onset patients is likely a more accurate indicator of possible liver dysfunction. This would be an important consideration for AAV9 gene therapy for GM1 patients, as patients in other rare disease cohorts treated with systemic AAV have experienced liver toxicity.^43^

Of course, the brain is anatomically, chemically, and functionally the focal point of pathology in patients with GM1. A previous study^27^ (including some of the current participants) indicated that quantitatively defined atrophy was more severe among participants with late infantile onset compared to juvenile onset. In this larger cohort, qualitative ratings indicated myelin disruptions or abnormalities in all late infantile and most juvenile onset patients, in addition to considerable atrophy. From the perspective of clinical trial readiness, it seems plausible that some intervention may slow atrophy, but a reversal seems unlikely. Other indicators of neuronal status may therefore be useful as biomarkers of disease severity and/or response to treatment.

As suggested by others^27^, MRS may be fit for this purpose. In the current study, the quantification by MRS of six metabolites in the left centrum semiovale documented significant excess of myo-inositol (a measure of gliosis) and deficits in NAA (a measure of neuronal health). These findings are consistent with the neuronal loss and increased gliosis observed in murine and feline models of GM1 disease.^44–47^ A strength of this study was our ability to compare these putative biomarkers with both age and behavior, finding that abnormalities in both myo-inositol and NAA were related to age and adaptive functioning. Future work is needed to elucidate their viability as biomarkers for type II GM1.

In summary, although variability in type II GM1 gangliosidosis disease progression is evident in both subtypes we show here that no child escapes a progressive downward trajectory. This work builds upon the existing literature by using direct multisystem assessments over a long period of time within person, and a wide age range across individuals. Given the variability in type II GM1 gangliosidosis it will be important to design outcome measures for therapeutic trials that are sensitive to stability or subtle improvement, realizing that some features (e.g., cerebral atrophy) will not be reversible. Early diagnosis and newborn screening are therefore critical in identifying pre-symptomatic patients who have the greatest chance of deriving benefit from mechanism-modifying therapy. Fortunately, a collaboration of academicians, industry, and patient advocacy groups is actively engaged in this endeavor.^48^

## Data Availability

The data described in this manuscript are available from the corresponding author upon reasonable request.

## Acknowledgments

We thank the participants and their families for the generosity of their time and efforts. We are also grateful to many staff members and care providers who contributed their expertise over the years.

## Author Contributions

Conceptualization: CT, JJ, PD, AT; Data curation: YA, PD, CF, RH, EN, CR, RQ, EB, MZ, SB; Formal analysis: CF; Funding Acquisition: AT, CT; Methodology: DA, EB, CB, BB, AB, STH, ALH, LAH, CJ, KK, JM, DR, BS, AT, CT, GB, WZ; Visualization: CF, CJ, DA, STH, WZ; Writing-original draft: PD, CF, CT; Writing-review & editing: YA, DA, EB, CB, BB, AB, AC, PD, CD, STH, ALH, RH, LAH, JJ, CJ, KK, JM, EN, RQ, DR, CR, BS, AT, CT, GV, MZ, SB, WZ

## Ethics Declaration

The NIH Institutional Review Board approved this protocol (02-HG-0107). Informed consent was completed with parents or legal guardians of the patients. All participants were assessed for their ability to provide assent; none were deemed capable.

## SUPPLEMENTARY METHODS

### Symptom onset and milestones

Parents responded to a study-specific questionnaire asking about the nature and timing of milestone attainment (i.e., sitting, crawling, walking, talking) and the nature and timing of the earliest symptoms. Data from this questionnaire were combined with semi-structured clinical interview data from the neurodevelopmental assessment (see below). Finally, we attempted recontact via phone with participants in August 2023 to update the onset of symptoms (e.g., gastronomy tube placement) and acquisition or loss of milestones. Recontact data from participants who had enrolled in NCT03952637 were not included, as this information reflected disease state after the administration of gene therapy.

### Ophthalmology

Eye exams were performed by pediatric ophthalmologists with ophthalmic genetics expertise. The ophthalmic assessment included a measurement of visual acuity with a Snellen chart or equivalent. When patient age, cognitive level, or cooperation precluded Snellen testing, Teller acuity was attempted to try and quantify visual acuity. The assessments of visual function are hierarchical, such that no blink to light reflects poor vision, followed by blink to light, occasional fix/follow, fix/follow, and central/steady/maintained or unmaintained. Achievement of the highest category indicates achievement of lower categories. Exams also included documentation of eyelid ptosis, ocular alignment, presence or absence of nystagmus, corneal and lenticular clarity, and fundus appearance including optic nerve head status. Where possible, clinical findings were supplemented with eye exams performed while the participant was under anesthesia for study related imaging procedures.

### Audiology

Audiological testing included age- and ability-appropriate behavioral assessments of hearing for pure-tone and speech stimuli, measures of middle ear function (tympanometry), and noninvasive physiologic testing (distortion product otoacoustic emissions) to assess cochlear health and supplement data from the behavioral assessment. For some participants, a neurodiagnostic auditory brainstem response (ABR) was performed; rarefaction and condensation click stimuli were presented separately at a high level (e.g., 85 dB nHL) with a low repetition rate (8.3 clicks/s). ABRs were interpreted using wave presence and morphology, absolute and interpeak latencies, and the wave V/I amplitude ratio.^1^ However, due to poor patient cooperation, complete behavioral audiogram assessments were available for only 11 (46%) juvenile participants and no late infantile participants.

### Video fluoroscopic assessments of swallowing (VFSS)

When clinically indicated from historical data review (e.g., historical and current indicators of possible dysphagia, including weight loss, recent bronchitis, aspiration pneumonia, complaints of swallowing difficulties with one or more food textures, coughing while drinking, throat clearing after drinking, and/or complaints of food becoming stuck within the throat), video fluoroscopic assessments of swallowing (VFSS) were conducted systematically, assessing liquid, puree, and solid textures. Swallowing function was interpreted for appropriate safe swallowing. Following each study, the speech language pathologist summarized physiologic findings and applied a modified version of the American Speech-Language-Hearing Association / National Outcomes Measurement System (ASHA/NOMS) Dysphagia Scale, Diet Restrictions, and Modification Ratings^2^ to rate the overall ability to swallow safely from 0 (most severe) to 5 (least severe). The extent of aspiration/laryngeal penetration was rated using a 0 (most severe) to 5 (least severe) ordinal scale adapted from the Rosenbek Penetration/Aspiration Scale^3^ (PAS). These scales are described in Supplementary Table S8.

### Abdominal ultrasound

Liver size was calculated using longitudinal dimensions (mm) of right lobe of liver versus height and age.

### Mobility assessment

Progressive loss of the ability to ambulate either by crawling (late infantile) or walking (juvenile) is a hallmark of Type II GM1 patients. Due to significant cognitive impairment in this population preventing cooperation with verbal instruction, existing scales of mobility were deemed inappropriate. Therefore, an upright mobility scale (juvenile onset only) and a floor mobility scale (late infantile only) were developed and used in this study. Each is an ordinal scale from 1 (patient unable to perform the skill) to 5 (normal mobility for age). The scales are printed in **Supplementary Table S8**.

### Beta-galactosidase enzyme assay

The activity of β-galactosidase was measured using a synthetic fluorogenic substrate as previously described.^3^ CSF samples were analyzed for βgal activity using 30µL of sample and serum was analyzed using 10µL of sample. For both CSF and serum, 100µL of substrate was added followed by incubation at 37°C for 1 hour. Protein concentration was determined by Lowry method, utilizing 30µL CSF or 1µL serum, and used to normalize βgal activity to nmol 4MU cleaved/mg protein/hour. Data is expressed as fold of normal and determined by dividing the specific activity of the sample by the average of specific activities from age-matched control samples. Floor effects in β-galactosidase prevent significant declines over time, and so when patients had multiple serum and/or CSF samples, their median value was used.

### Electroencephalogram

A 21-channel digital EEG with time locked video and single-lead EKG was performed. EEG electrodes were placed according to the international 10-20 system of electrode placement. The EEG was reviewed by a board-certified pediatric epileptologist (blinded to patient group assignment) using the longitudinal bipolar montage (and when appropriate, referential and transverse montages). Intermittent photic stimulation and hyperventilation were performed in studies that were <1 hour in duration. The total duration of recording ranged from 20 minutes to 12 hours.

A small number of patients also underwent polysomnography (PSG). A one-night, clinical PSG was conducted either on the pediatric ward with a portable system or in the sleep lab at Clinical Center at NIH. Each study included overnight monitoring of electroencephalogram, electro-oculogram, ECG, chin and anterior tibial electromyogram, nasal pressure transducer, oral thermistor, a snore sensor, respiratory inductive plethysmography, pulse oximetry and continuous video monitoring. Studies were attended by a certified sleep technologist experienced in pediatrics. Studies were manually scored according to the American Academy of Sleep Medicine Manual version 3, for the scoring of sleep and associated events. The study reports were assessed and approved by a pediatric sleep physician. Given the small number of participants, these results are found in **Supplementary Table S7**.

### Magnetic resonance imaging

Brain MRI studies conducted at the NIH Clinical Center were all performed on the same 3T Philips scanner, using an 8-channel SENSE (sensitivity encoding) head coil. Deep sedation with monitored anesthesia care or general anesthesia was used for all participants. Clinical MRI examination included sagittal T1-weighted, axial T2-weighted, coronal STIR, axial fluid attenuated inversion recovery (FLAIR), diffusion tensor, and 3D MP-RAGE (magnetization-prepared rapid acquisition with gradient echo) images, without intravenous contrast. For a subset of participants, MRI studies were obtained from the subjects’ referring institution for comparison. All MRI scans were reviewed by the same pediatric neuroradiologist (GV).

Qualitative ratings were made based on the following information. The size of the brainstem was assessed on the mid sagittal T1-weighted images, assessing the surface area of the midbrain, pons and medulla. The cross-sectional area of the corpus callosum was assessed qualitatively on mid sagittal T1-weighted images; for infants, the degree of callosal atrophy was estimated based on age-expected size. The degree of cerebellar atrophy was estimated based on the size of the cerebellar folia. The degree of cortical atrophy was estimated based primarily on the size of the cerebral cortical sulci, also size of the lateral ventricles. For infants, white matter abnormalities were defined primarily as a delay in myelination compared to age expectations, as demyelination/gliosis is not easily assessed in this age group. After infancy, when myelination is essentially complete, white matter abnormalities were assessed based on the extent and progression of abnormal increased signal on T2-weighted images. The abnormal increased signal is representative of either demyelination, gliosis or dysmyelination, often in combination. The size and signal intensities of the basal ganglia and of the thalami were also evaluated.

### Magnetic resonance spectroscopy

Quantitative single-voxel 1H-MRS was performed on three locations; here we report only on the left centrum semiovale (LCSO). Voxels were graphically prescribed from sagittally-acquired 3D-TFE images reformatted into three planes. 1H-MRS acquisition was performed with PRESS localization, CHESS water suppression, TE=38 ms, TR=2000 ms, and NEX=128. An unsuppressed water spectrum (TR=5000 ms, TE=38 ms, NEX=16) was also acquired for each voxel. Identical gain and shim settings were used for both spectra from each voxel so that metabolite concentrations could be determined. To correct for CSF included within the voxels, we acquired a heavily T2-weighted image with location and slice thickness corresponding to the location of each MRS voxel (FSE; ETL=8; TE=500 ms; TR=3000 ms), and a phantom containing water was placed beside the head and included in the field-of-view of the CSF correction image.

Post-processing of the spectra was performed using LCModel^4^, followed by correction for estimated tissue water^5^ and T1 of the metabolites within tissue ^6^. Because only one echo time was acquired, no correction was made for T2 decay of the metabolites. We analyzed creatine, myo-inositol, choline-containing compounds (“choline”), *N*-acetylaspartate+*N*-acetylaspartyl glutamate (“NAA”), and glutamine+glutamate+gamma-aminobutyric acid (“Glx”). Post-processing for correction of CSF partial volume was done as described in other reports^7–9^.

We generated reference curves for each metabolite using data from 38 individuals aged 1-42 years (interquartile range: 3.6–23.3 years). Similarly-acquired and post-processed data from healthy children are not available; instead, children in the reference group were asymptomatic or neurologically pre-symptomatic participants in other protocols at our institution who were scanned and post-processed with the same method as the GM1 participants. Adults in the reference group were healthy volunteers. The reference curve for each metabolite was generated by fitting a model of the form y=Ax^B^ (where x is age and y is metabolite concentration) to the reference data. The difference from the reference curve value was used in all analyses.

### Neurodevelopmental Assessments

Speech and language assessments were conducted utilizing informal play and conversation speech tasks involving auditory comprehension and speech production, which were rated by a single SLP on an observational outcome matrix for speech and language functions that also ranged 0 (most severe) to 5 (least severe). Each of these rating scales is reproduced in **Supplementary Table S6**.

Adaptive behavior is the collection of conceptual, social, and practical skills a person uses to function in everyday life. Adaptive behavior is a primary behavioral indicator of the effects of disease progression, and this high degree of clinical relevance is why it was selected for inclusion. The Vineland Adaptive Behavior Scale comprehensive semi-structured parent/caregiver interview is widely used to measure adaptive behavior within the context of neurodevelopmental disorders, and it is appropriate for the full age range (birth to 99 years). Here we began with the version available at study initiation (Vineland-II^10^) and added the current version (Vineland-3^11^) when it became available and its improved psychometric profile was demonstrated.^12^ The Vineland was administered by experienced clinicians in a semi-structured interview with caregivers. The Vineland yields an overall composite and domain-level standard scores (mean = 100, SD = 15), and V-scale (mean = 15, SD = 3) and growth scale value (GSV; Vineland-3 only) scores at the subdomain level. GSVs are transformed raw scores that are intended to measure change over time in an individual. GSVs are particularly helpful in detecting stability or subtle improvements over time in individuals with significant levels of impairment at baseline.^13,14^

### Statistical models for longitudinal data

For outcomes with sufficient numbers of repeated measures, formal longitudinal modeling was attempted, using the following methods. The timescale of interest in this study was chronological age, which was decomposed into between-subject (average chronological age during the study period) and within-subject (duration of participation) effects. While the between-subject effect is useful to understand the developmental course, the within-subject effect more directly corresponds to change that can be expected during a time period relevant to clinical trials (e.g., yearly change). Uncorrelated random effects were used to account for repeated measures within individual (subject-level intercept and slope of within-subject time). While participants were also technically clustered within site and family, the size of the datasets did not support the complexity of modeling this clustering and it was ignored. The parameters of interest from the growth models were the intercept and slope estimates with 95% confidence intervals. Model assumptions were evaluated graphically.

## SUPPLEMENTARY RESULTS

### Medication use

Participants continued routine clinical care at their home institutions, and medication compliance and its effect on disease was not measured. At their initial visit, eleven (65%) late infantile patients and seven (29%) juvenile patients reported treatment for seizures. There are no FDA-approved medications for the treatment of GM1 gangliosidosis, however off-label use of substrate inhibitor, miglustat, and amino-acid analog, *N*-acetyl leucine are common in GM1 and GM2 patients. During study participation, nine (38%) juvenile and four (24%) late infantile patients took miglustat, and two (8%) juvenile and two (12%) late infantile took *N*-acetyl leucine.

### Growth

Head circumference was within the normal range at the first available assessment, the median [IQR] age- and sex-based percentile was 58 [33.3, 89] in the late infantile group and 40.5 [6.3, 85] in the juvenile group. BMI was also generally within the normal range at the first available assessment. The median [IQR] age- and sex-based Z-score was - 0.11 [−0.85, 0.75] in the late infantile group and 0.11 [−1.10, 0.59] in the juvenile group (**Supplementary Figure S8**).

## SUPPLEMENTARY TABLES & FIGURES

**Table S1. Mutation classifications.** The table shows the evidence and grading scheme used for single nucleotide substitutions. Two deletions (exon 3-5 deletion and exon 5 deletion) not shown in the table are out of frame and thus are pathogenic. Categorizations: Very Strong (PVS), 8 points; Strong (PS), 4 points; Moderate (PM), 2 points; Supporting (PP), 1 point. Classifications: Pathogenic (PATH), 10+ points; Likely Pathogenic (LPATH), 6 to 9 points; Variant of Uncertain Significance (VUS), fewer than 5 points. Cumulative count of classifications: PATH, 28; LPATH, 7; VUS, 0.

*See separate file*.

**Table S2.**
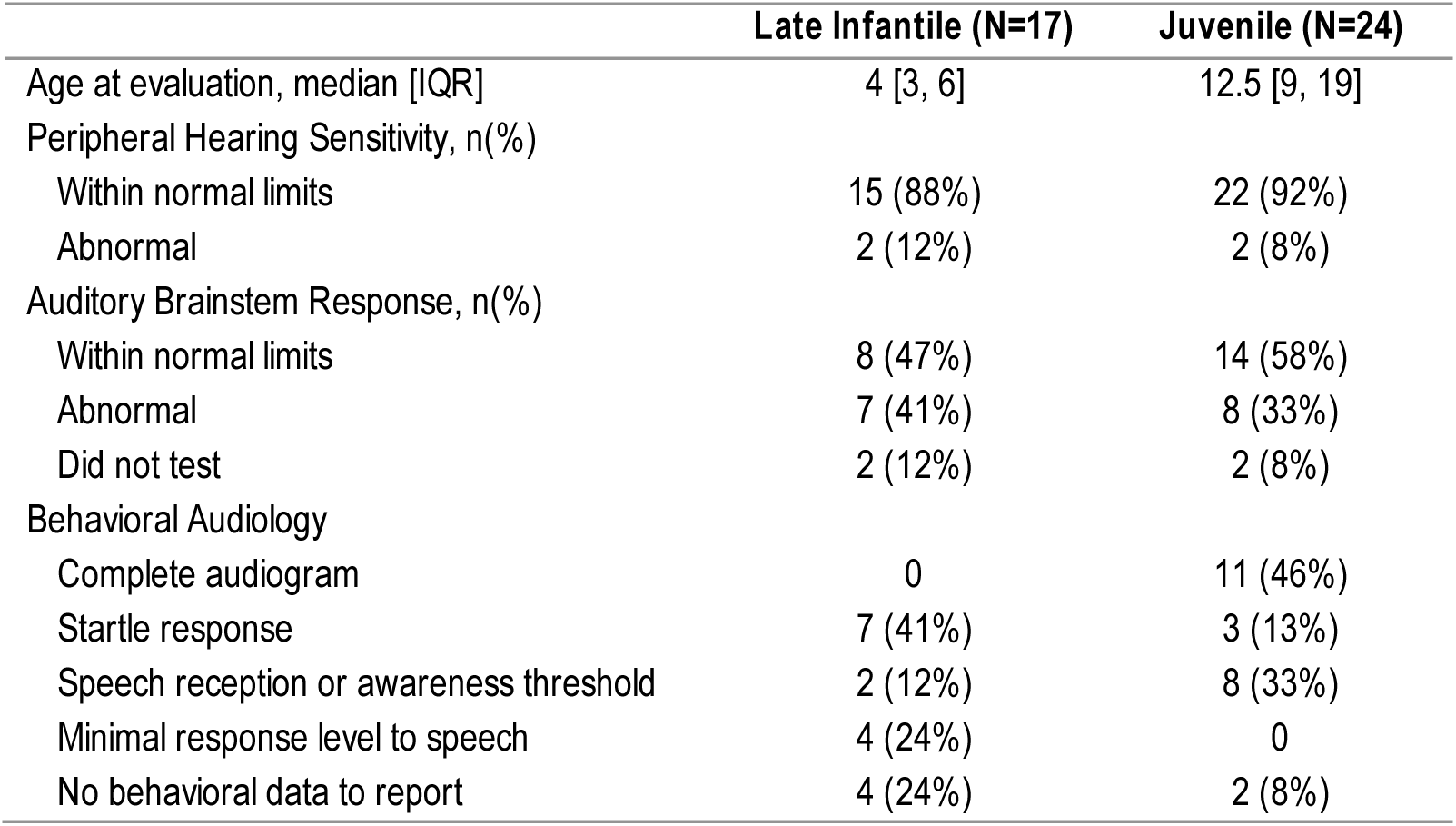
Audiology Results at Reference Evaluation. Note: Reference evaluation was the most recent, most complete evaluation for each participant. Auditory brainstem response was completed under sedation for 10 (of 15, 67%) of the late infantile onset group and 14 (of 22, 64%) of the juvenile onset group.

**Table S3.** Liver enzymes. Data fromthe earliest available observationper personwas used (N = 17 late infantile and N=24 juvenile). The distribution of age in years (median [IQR]) was 4.79 [3.53, 5.68] for the Late Infantile group and 11.82 [7.56, 14.65] for the juvenile onset group.

|  | Late Infantile (N=17) |  |  | Juvenile (N=24) |  |  |
| --- | --- | --- | --- | --- | --- | --- |
| | *Median [IQR]* | *n (%)*<br>*abnormally*<br>*high* | $\rho_{age}$ | *Median [IQR]* | *n (%)*<br>*abnormally*<br>*high* | $\rho_{age}$ |
| AST, U/L | 73 [44.35, 93.85] | 13 (76) | -0.42 | 33.84 [27.15, 40.47] | 7 (29) | -0.53 |
| ALT, U/L | 20.8 [16.6, 27] | 3 (18) | -0.42 | 18.7 [16.42, 25] | 2 (8) | 0.17 |
| GGT, U/L | 13.1 [12.11, 26.6] | 6 (35) | -0.05 | 13.8 [8.77, 22.18] | 3 (13) | 0.1 |

**Table S4a.** Summarystatistics for cross-sectional MRS data. MRS concentrations (mM) are expressed as deviations from age expectations. Spearman correlations were calculated between difference score and age. The cross-sectional dataset containedthe earliest available observationper person. Age at assessment (M[IQR]): Juvenile, 11.66 [7.47, 13.56]; Late infantile, 3.82 [3.43, 5.94].

|  | Juvenile (n=21) |  | Late infantile (n=13) |  |
| --- | --- | --- | --- | --- |
| | Median [IQR] | $\rho_{age}$ | Median [IQR] | $\rho_{age}$ |
| Creatine difference, mM | 0.26 [-0.86, 0.64] | 0.22 | 0.69 [-0.51, 0.85] | -0.57 |
| Myo-inositol difference, mM | 1.12 [0.74, 2.18] | 0.68 | 1.6 [0.37, 1.97] | 0.28 |
| Choline difference, mM | 0.08 [-0.13, 0.17] | -0.31 | 0.03 [-0.43, 0.23] | -0.46 |
| NAA difference, mM | -2.13 [-4.01, -0.3] | -0.74 | -3.45 [-6.81, -0.57] | -0.59 |
| Glx difference, mM | 0.21 [-0.05, 1.28] | -0.51 | -0.49 [-1.62, 0.17] | -0.15 |

**Table S4b.**
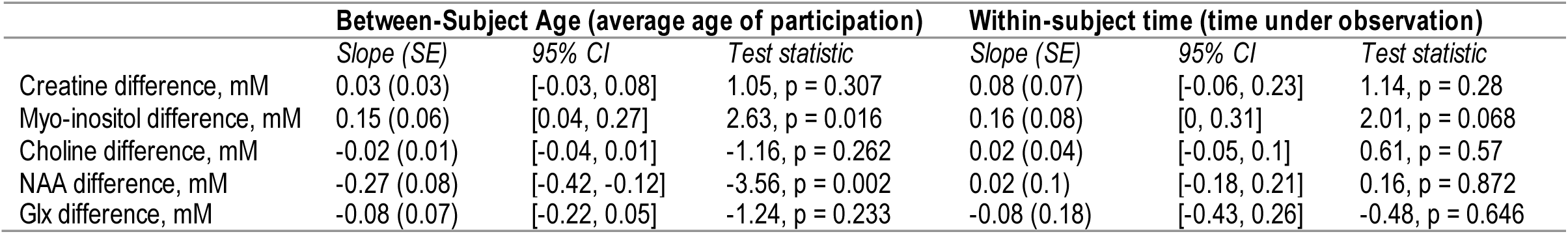
Juvenile cohort: Parameter estimates and test statistics from mixed effects model of age- and time-related trends in MRS concentration (mM, difference from age expectations). The interpretation of between-subject age is the expected difference in MRS concentration between two people with a difference in average age of participation of 1 year. For example, older participants had higher myo-inositol concentration differences relative to younger participants, at a rate of 0.15 per additional year of age. The interpretation of within-subject time is the expected change within an individual person over the course of 1 year. For example, the average participant had an increase of myoinositol difference of 0.16 mM per year.

**Table S5.**
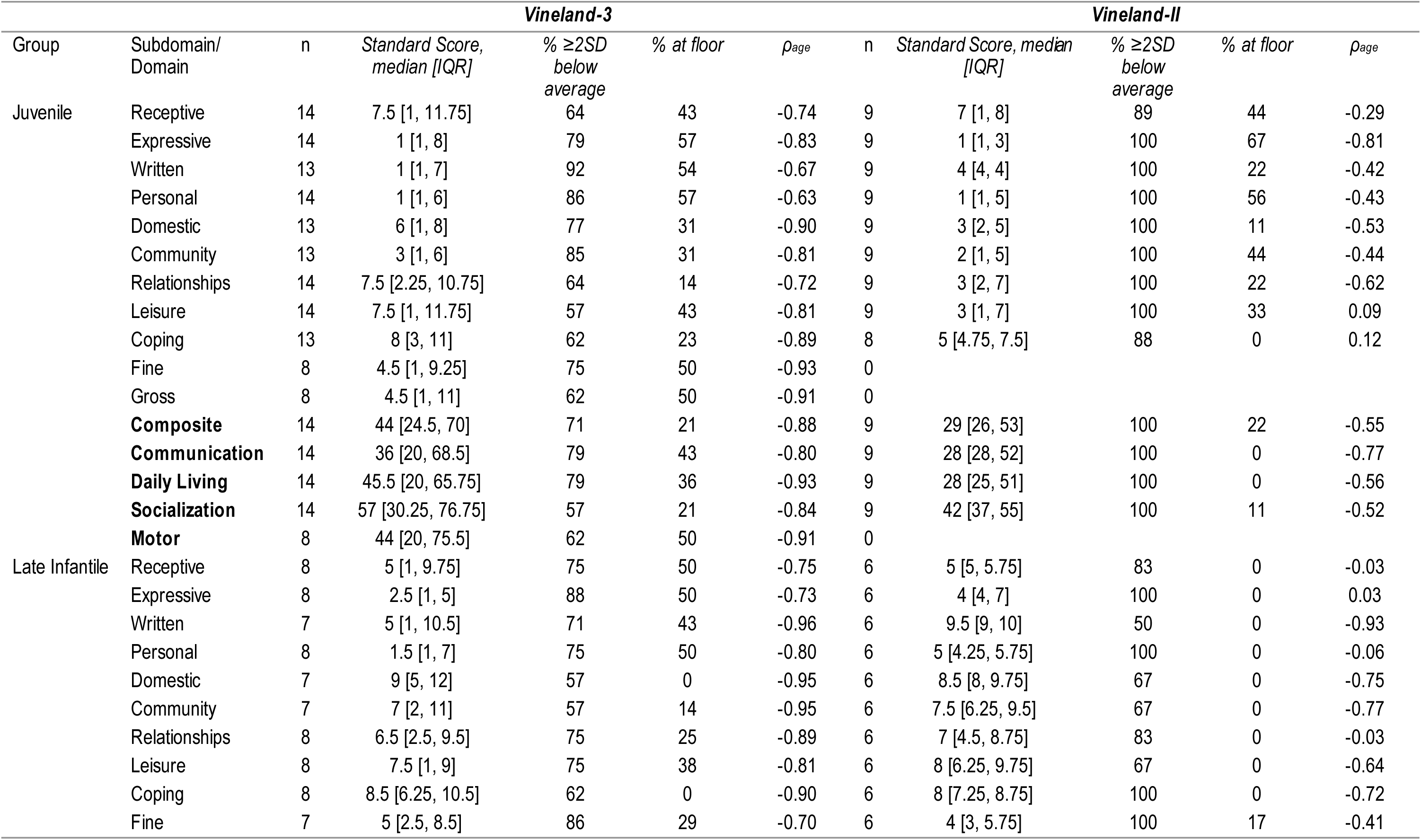
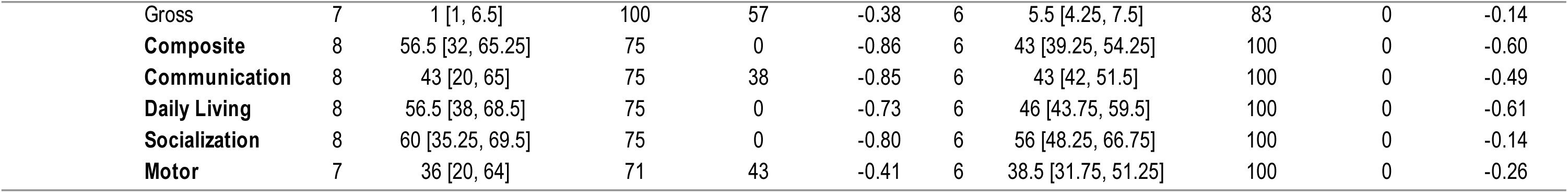
Subdomain-level V-scale summary statistics for the earliest assessment available for each participant. The first Vineland-II and/or first Vineland-3 assessment available for each person is summarized in this table. Sample sizes and ages were: Late Infantile/Vineland-3: n = 8 (median age: 3.97 [3.06, 5.98]), Late Infantile/Vineland-II: n = 6 (median age: 4.09 [3.46, 4.91]); Juvenile/Vineland-3: n = 14 (median age: 9.59 [7.37, 17.39]), Juvenile/Vineland-II: n = 9 (median age: 17.29 [11.99, 19.38]). Motor excluded as standard scores are not available for all participants. Receptive, Expressive, and Written subdomains belong to Communication; Personal, Domestic, and Community belong to Daily Living; (Interpersonal) Relationships, (Play and) Leisure, and Coping belong to Socialization. Standard scores have a population mean of 100 and SD of 15 (floor = 20), subdomain-level V-scale scores have a population mean of 15 and SD of 3 (floor = 1). IQR = interquartile range (25^th^ – 75^th^ percentile).

**Table S6.**
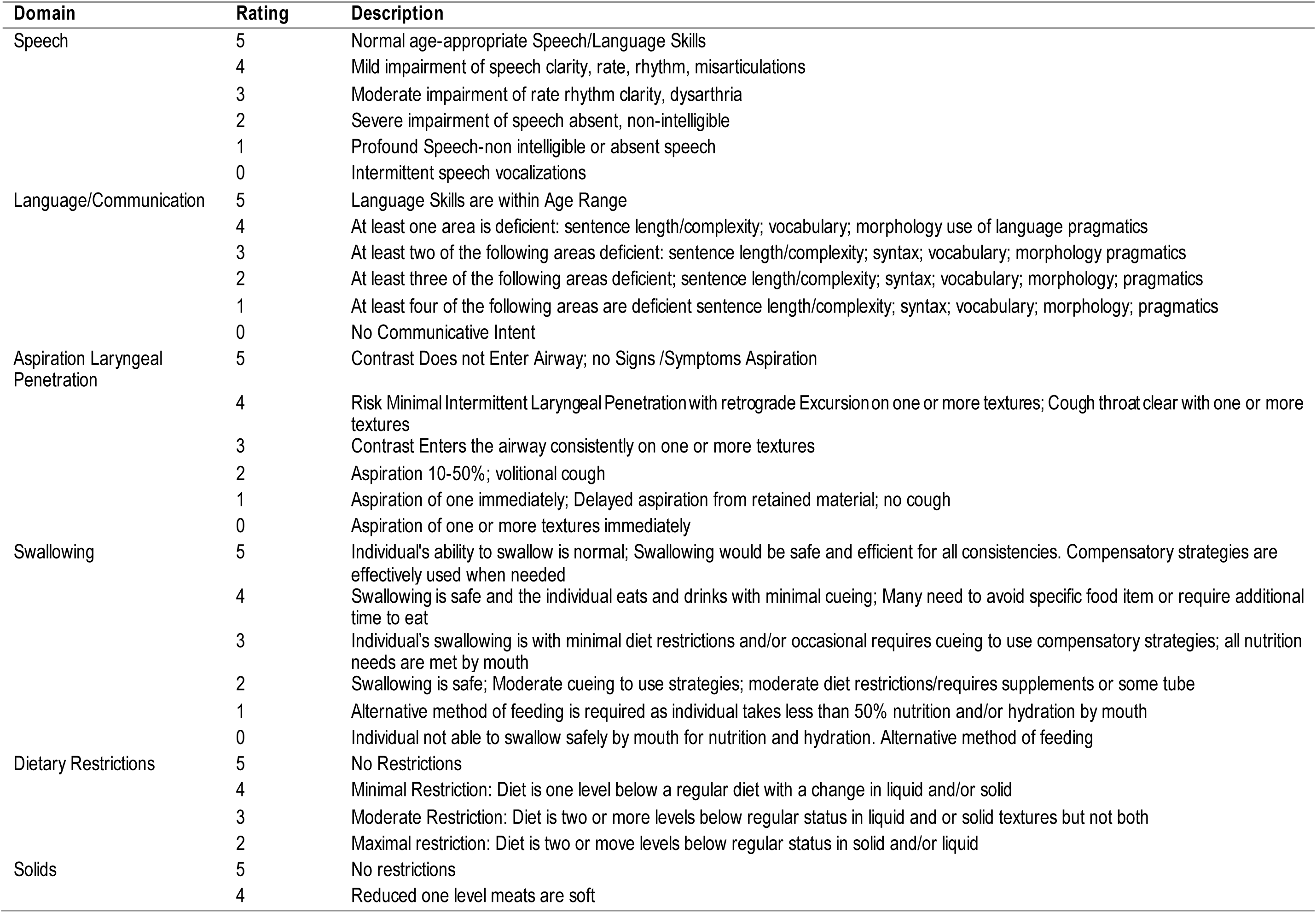
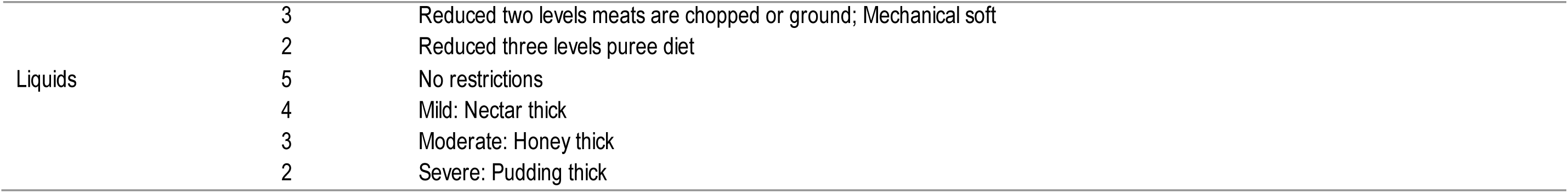
Scoring scales used for outcome measures of speech, language, aspiration/laryngeal penetration, swallowing, and dietary restrictions, partially adapted from ASHA/NOMs and Rosenbek scales.

**Table S7.** Polysomnography results for seven participants. LI = late infantile, JUV = juvenile, BMI = body mass index, AHI = apnea-hypopnea index, SWS = slow wave sleep, REM = rapid eye movement, WASO = wake after sleep onset

| Group | Sex | Age Range (years) | BMI | Pedi-Epworth Score | Recording Time (mins) | Total Sleep Time (mins) | Sleep Efficiency | Overall AHI (per hr) | Sleep Onset Latency (mins) | N2 Latency (mins) | SWS Time (mins) | REM Latency (mins) | REM Time (mins) | REM % of TST | WASO (mins) |
| --- | --- | --- | --- | --- | --- | --- | --- | --- | --- | --- | --- | --- | --- | --- | --- |
| LI | Male | 5 – 9 | 16.2 | 14 | 606 | 358 | 59.20% | 0.5 | 125 | 129 | 0 | 171 | 51 | 14.20% | 120 |
| LI | Male | 5 – 9 | 16.1 | 11 | 649 | 487 | 75.10% | 1.5 | 5 | 8 | 0 | 179 | 61 | 12.50% | 156 |
| LI | Female | 0 – 4 | 18.5 | 4 | 514 | 451 | 87.70% | 0.3 | 20 | 23 | 227.5 | 95 | 87 | 19.30% | 43.5 |
| JUV | Male | 30 – 34 | 19.8 | 5 | 558 | 335 | 60.10% | 0.9 | 93 | 103 | 0 | 179 | 17 | 5.10% | 128.5 |
| JUV | Female | 20 – 24 | 23.7 | 0 | 437 | 41 | 9.40% | 1.5 | 217 | 220 | 0 | 0 | 0 | 0% | 173.5 |
| JUV | Female | 15 – 19 | 17.4 | 6 | 666 | 569 | 85.40% | 0.5 | 3 | 11 | 0 | 64 | 180.5 | 31.70% | 94 |
| JUV | Male | 5 – 9 | 16.4 | 3 | 522 | 468 | 89.80% | 1 | 0 | 0 | 120.5 | 205 | 84 | 17.90% | 52.5 |

**Table S8.** Rating anchors for the mobility assessment. The floor mobility scale was used for the late infantile group and the upright mobility scale was used for the juvenile onset group.

| Rating | Floor Mobility | Upright Mobility |
| --- | --- | --- |
| 5 | Crawls in 4-point independently | Ambulation is normal for age |
| 4 | Scoots independently when sitting | Independent ambulation, may be unsteady |
| 3 | Sits without support | Ambulates independently with mobility aid due to unsteady gait |
| 2 | Sits with support | Ambulates only with the assistance of another person (with or without a mobility aid) due to unsteady gait |
| 1 | Rolls independently – rolling belly to back | Able to pull to stand, unable to ambulate |
| 0 | Unable to roll | Non-ambulatory, dependent for wheeled mobility (WC or stroller) |

**Figure S1.**
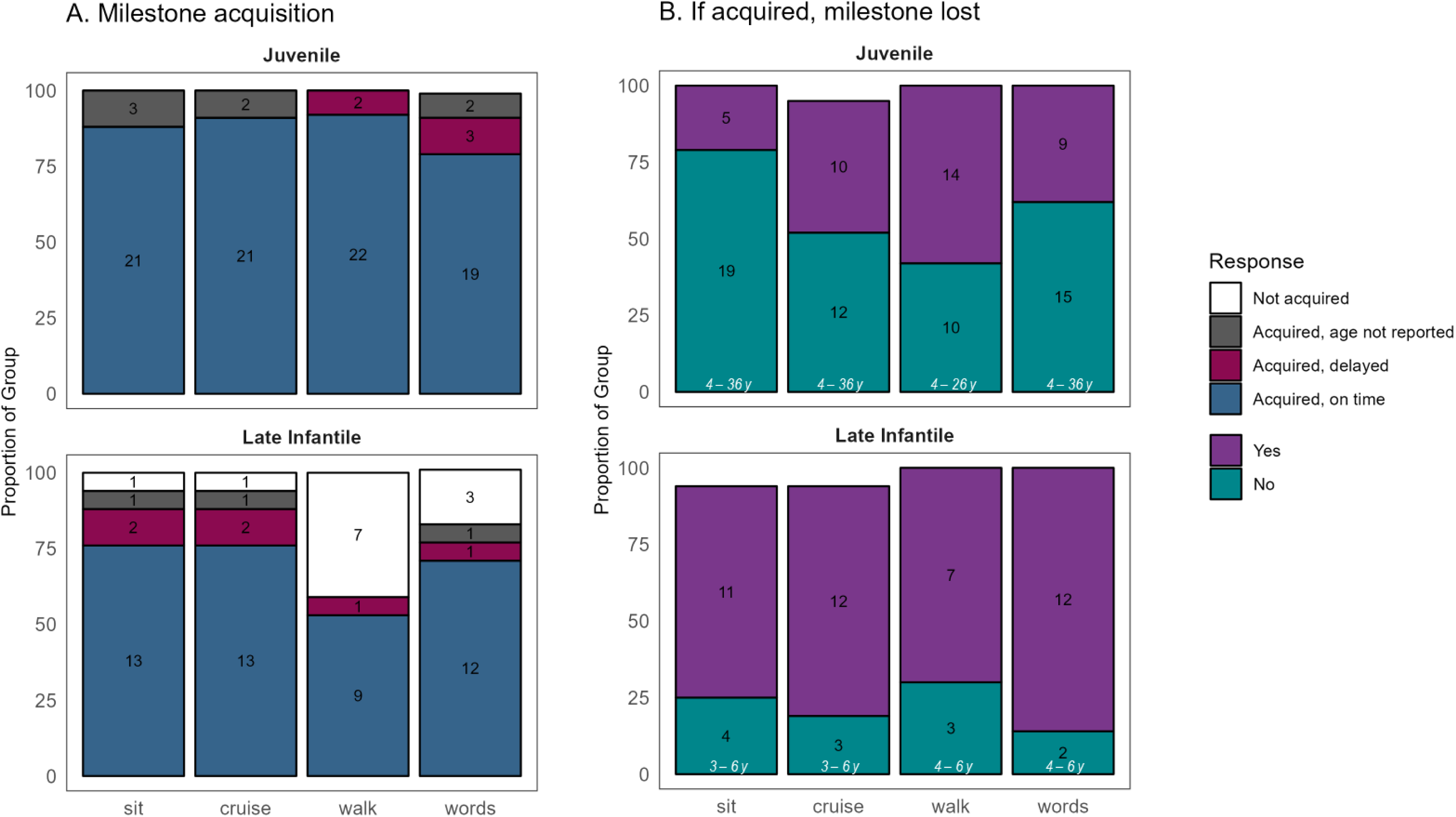
Parent-reported acquisition and loss of major milestones. Parent report of acquisition and loss of milestones was obtained at the last contact (for most participants, this was August 2023, see Figure 1). For Late Infantile, the age when these data were obtained/verified ranged 2.7 – 22.0 years, (median = 6.1, IQR = 4.2 – 11.2); for Juvenile 3.8 – 35.5 years, (median = 21.1, IQR = 14.2 – 26.1). Age of acquisition was coded as “on time” or “delayed” according to WHO or CDC normative data (thresholds: sitting: 8 months; cruising: 14 months; walking: 16 months; words = 12 months). If the participant acquired a skill, loss was queried. The number of participants in each category is inset in black text, and in panel B, the age range of those who had not yet lost the skill is inset in white text.

**Figure S2.**
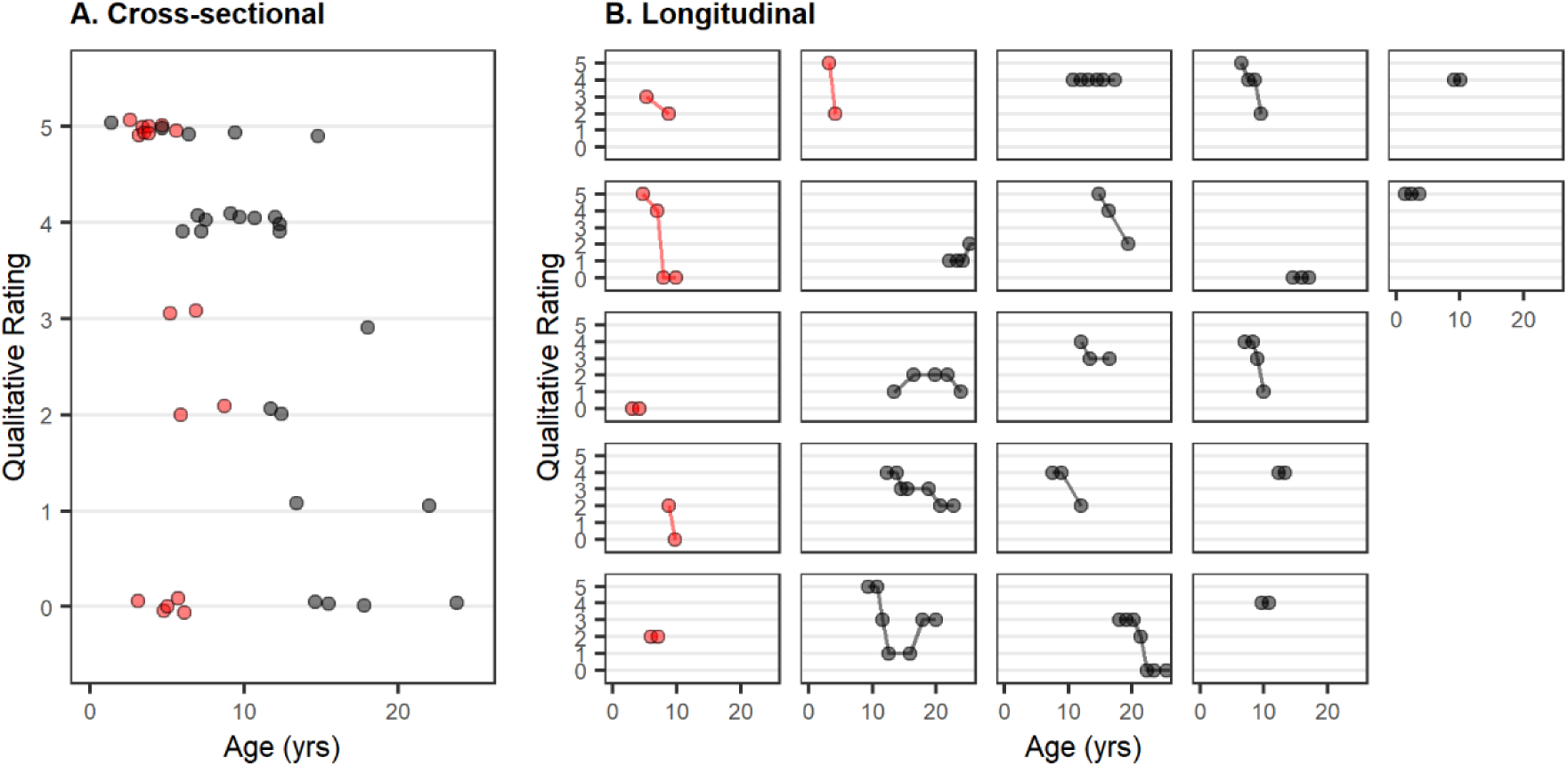
Floor (late infantile, red) and upright (juvenile, black) mobility scale ratings at baseline (Panel A) and across repeated measures(Panel B). Note that the qualitative ratings made on a scale of 0 (most impaired) to 5 (normal) differ between late infantile participants (red points; floor mobility) and juvenile participants (black points; upright mobility). See supplementary tables for qualitative rating descriptions. Slight random jitter added to Y-axis of cross-sectional panel (panel A) to reduce overplotting.

**Figure S3.**
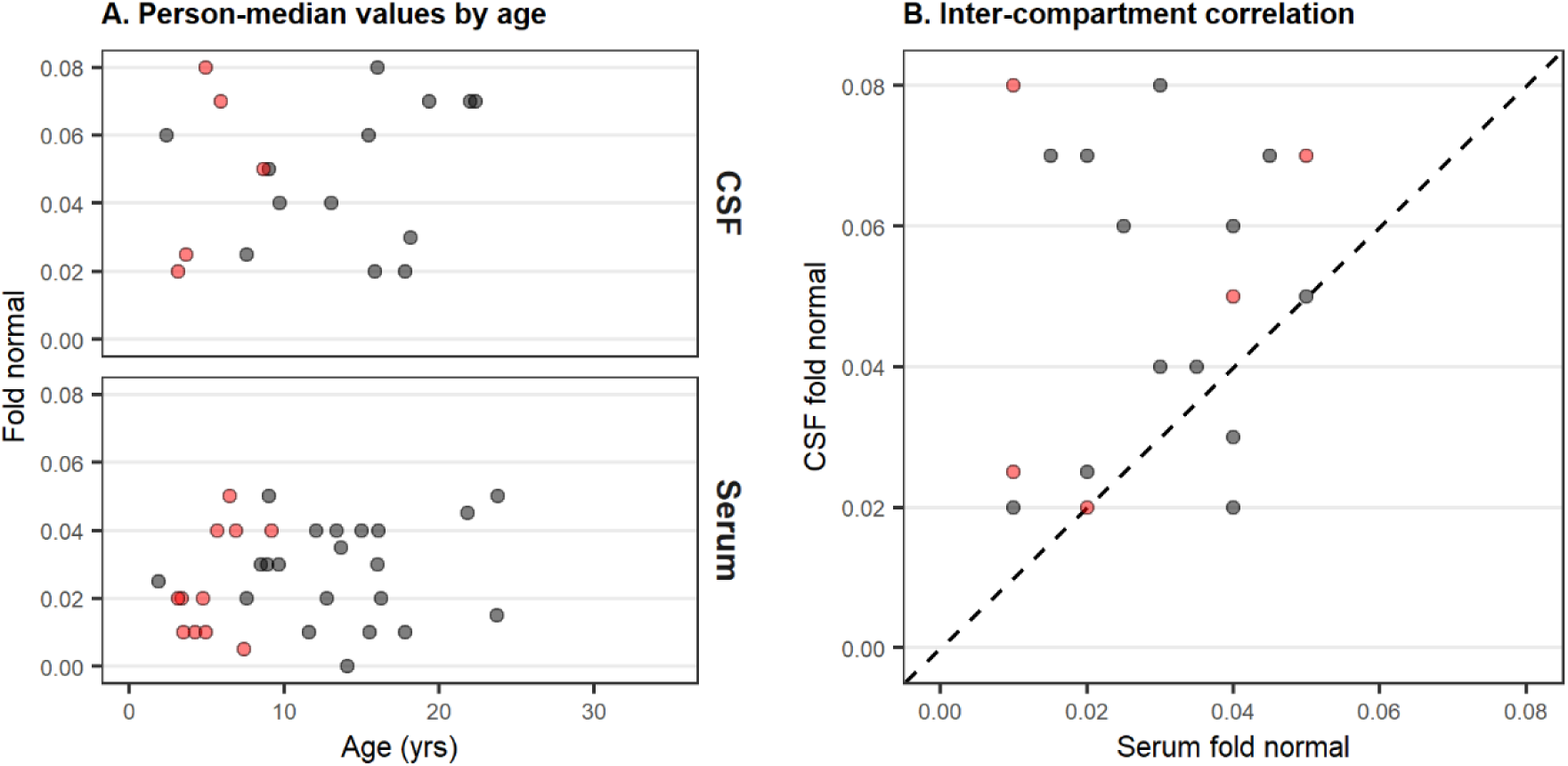
β-galactosidase enzyme activity in CSF and serum. Panel A: Fold-change from pediatric control sample. Panel B: Related to lack of variability in CSF and serum values, no correlation between CSF and serum was observed (Late infantile: ρ = 0.2; Juvenile: ρ = .04). Dotted line indicates 1:1 correlation. Both panels: red = late infantile, black = juvenile; median b-gal value per person.

**S4.**
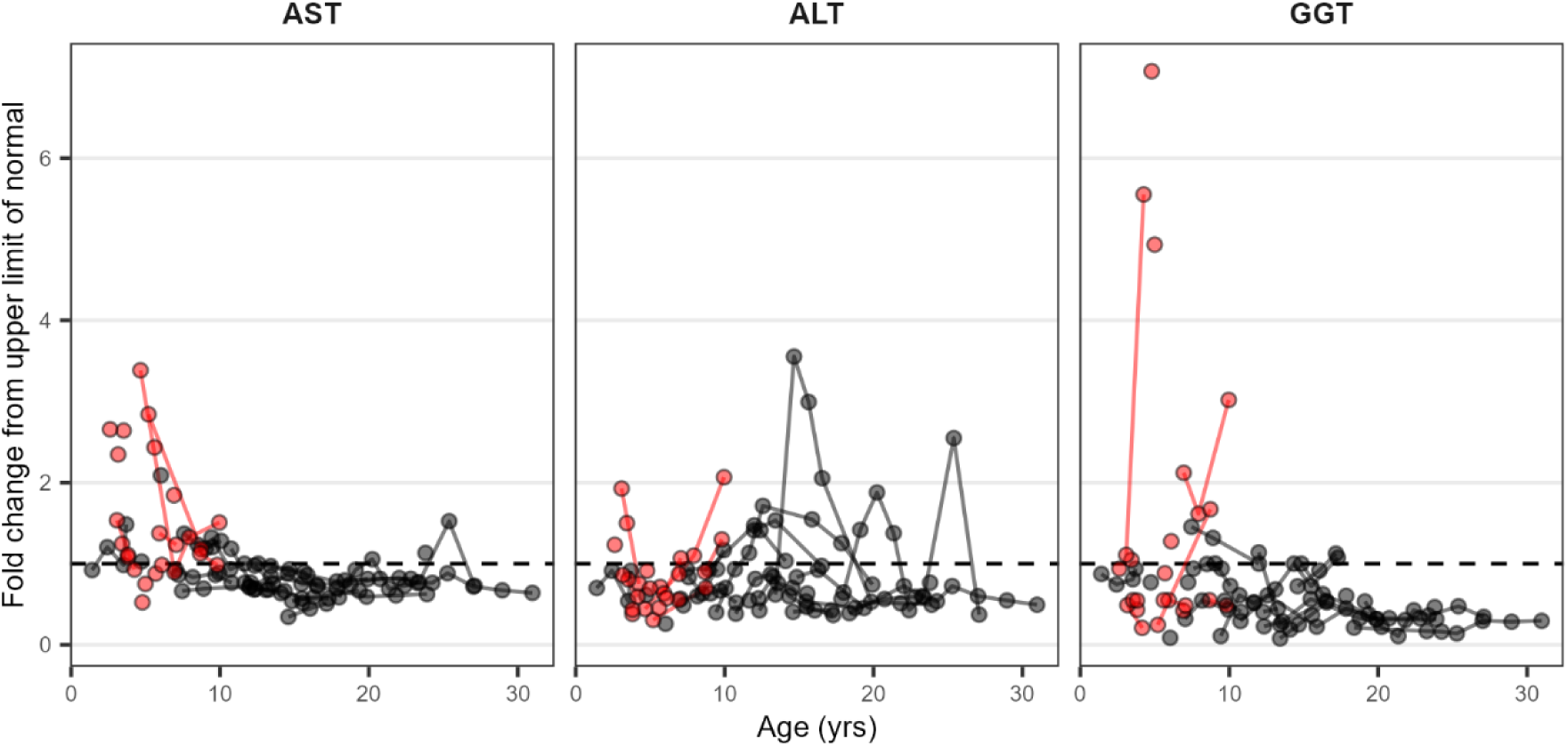
Longitudinal data on liver enzymes for late infantile (red) and juvenile (black) onset groups. Available data for each participant are plotted against their age and connected by a solid line. Concentrations were compared to reference range values and expressed as fold change from the upper limit of the reference range (dotted reference line at 1 indicates the upper limit of the reference range). Lines connect observations from a single person.

**Figure S5.**
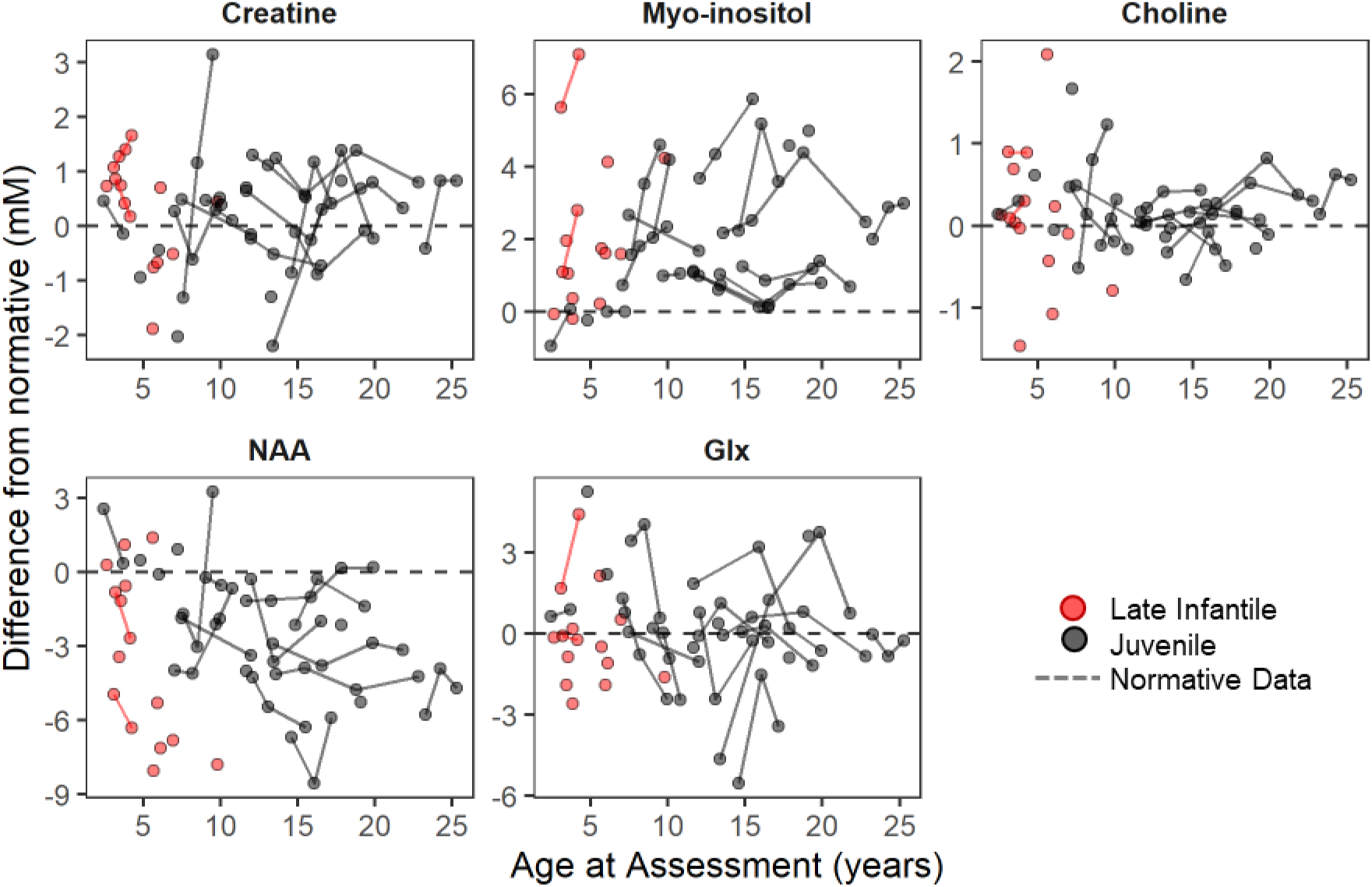
Longitudinal data showing age progression of metabolite concentration (mM) in LCSO relative to normative expectations. NAA: N-acetyl aspartic acid. Glx: Glu+Gln+GABA. Red = late infantile onset; black = juvenile onset. Lines connect observations from the same participant. Points show difference in concentration (mM) from age expectation, which is illustrated by the dotted line at zero.

**Figure S6a.**
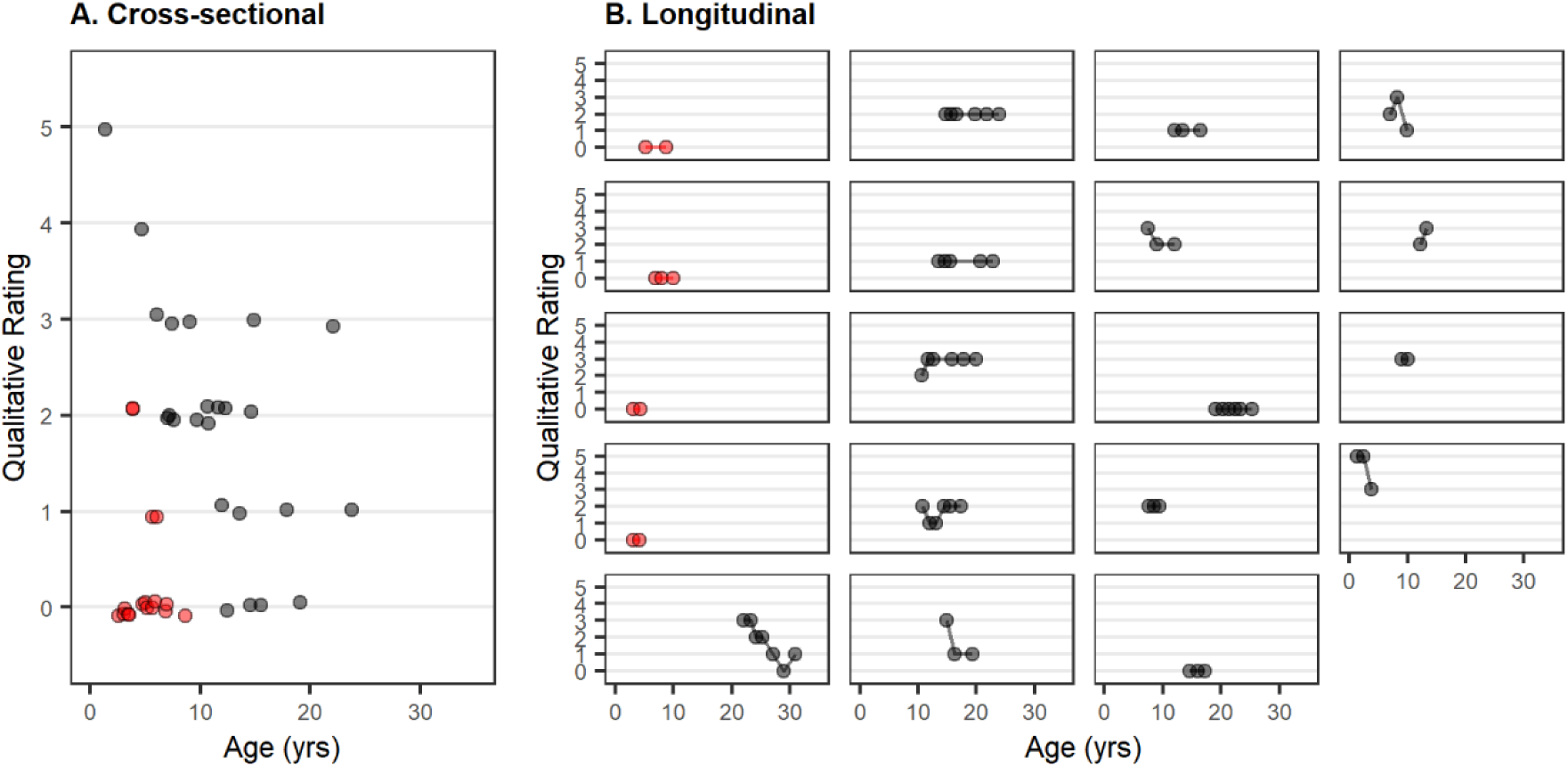
Speech qualitative ratings at first observation (panel A) and across longitudinal observation (panel B) for late infantile (red) and juvenile (black) groups. Qualitative ratings made on a scale of 0 (most impaired) to 5 (normal for age); see supplementary table for full description. Slight random jitter added to Y-axis of cross-sectional panel (panel A) to reduce overplotting.

**Figure S6b.**
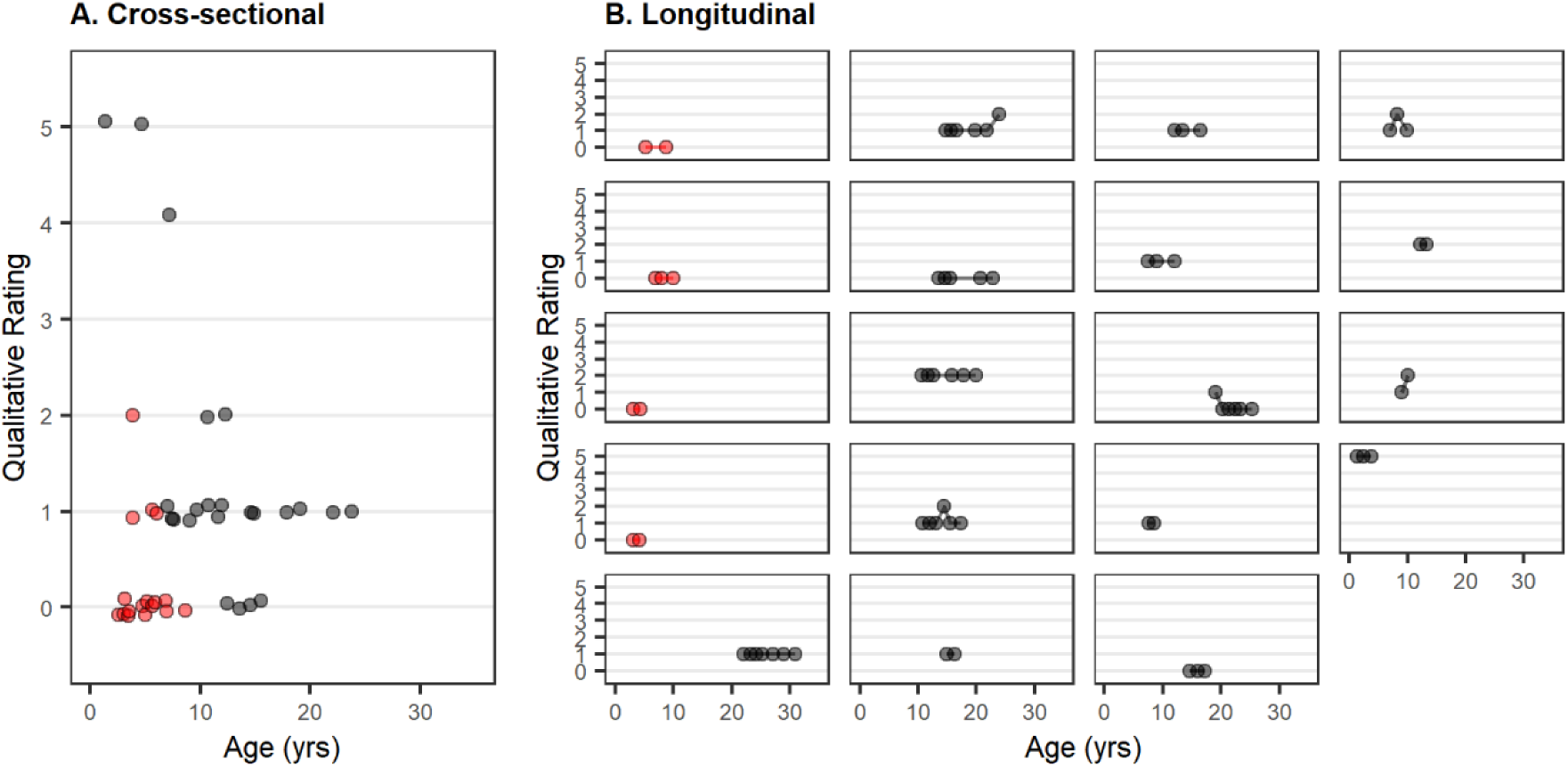
Language qualitative ratings at first observation (panel A) and acrosslongitudinalobservation (panel B) for late infantile (red) and juvenile (black) groups. Qualitative ratings made on a scale of 0 (most impaired) to 5 (normal for age); see supplementary table for full description. Slight random jitter added to Y-axis of cross-sectional panel (panel A) to reduce overplotting.

**Figure S7a.**
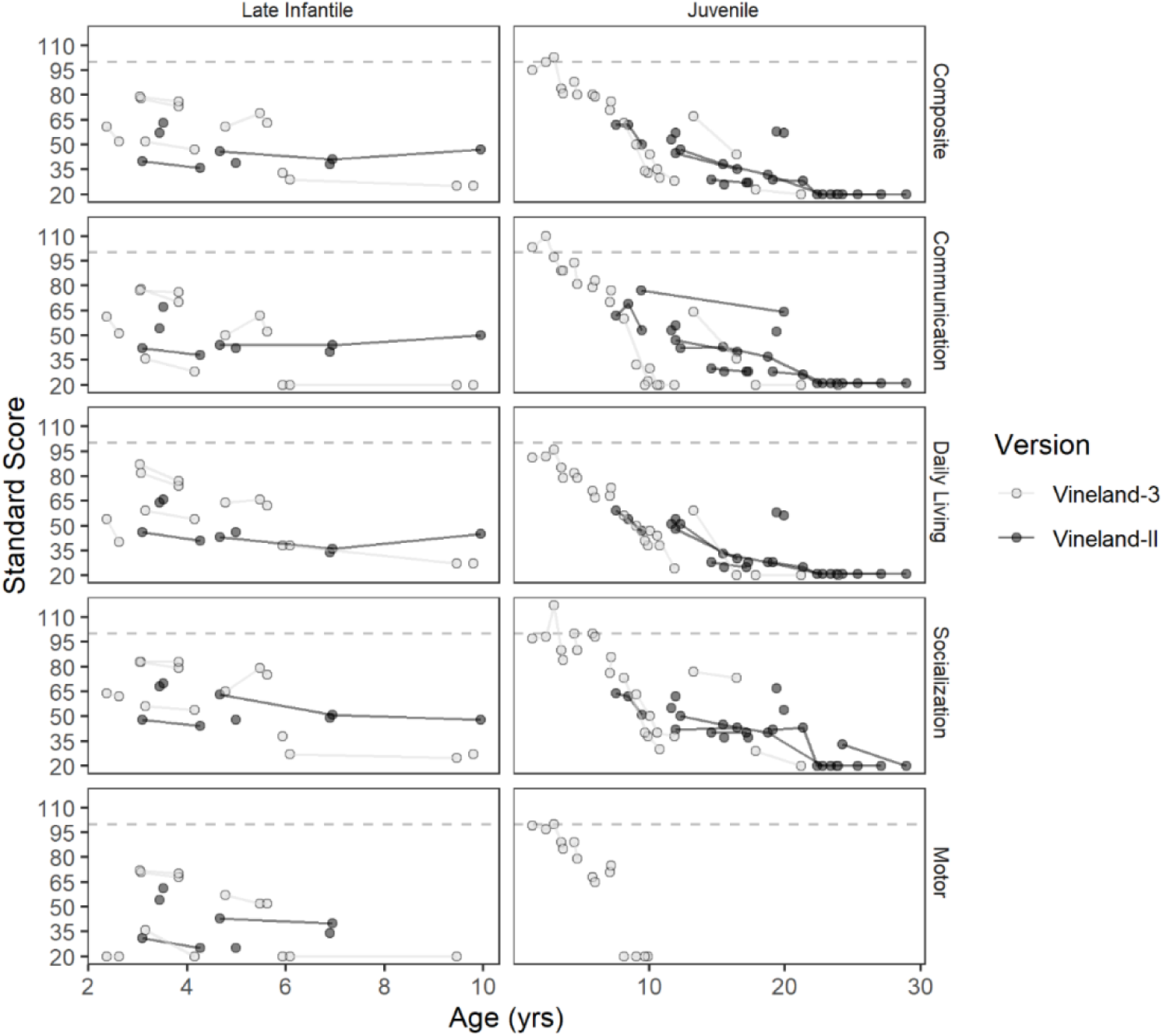
Longitudinal trends in Vineland domain-level standard scores for Late Infantile (left) and Juvenile onset (right). For each participant, the form (Vineland-II or Vineland-3) with a longer follow-up period was selected, so that each participant appears only once per panel. Where Vineland-II scores were used, the markers are black, and the markers are gray for Vineland-3. Solid lines connect observations from the same participant, but different participants are not otherwise demarcated. Dotted lines indicate normative expectations (population mean = 100, SD = 15).

**Figure S7b.**
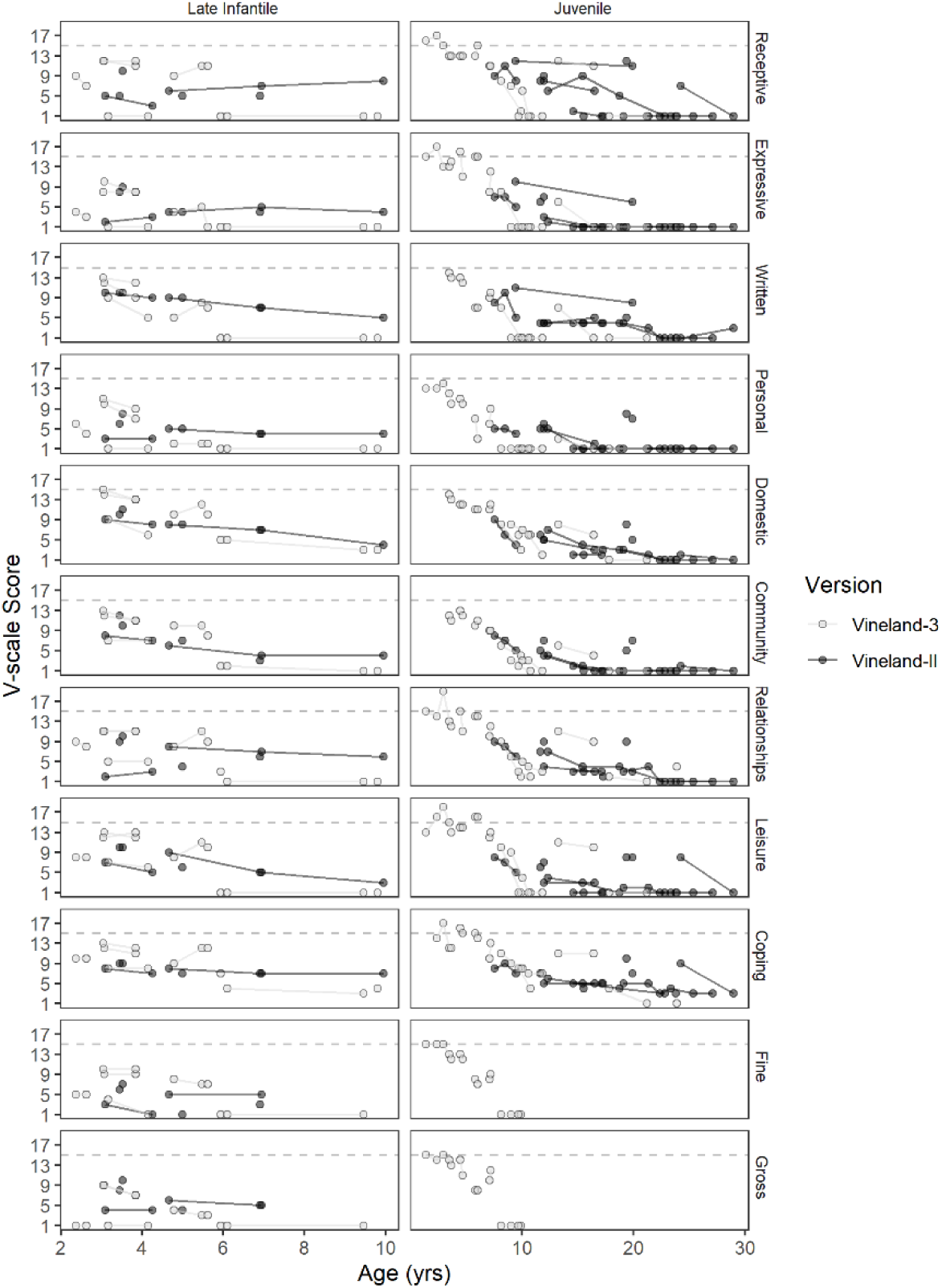
Longitudinal trends in Vineland subdomain-level V-scalescoresfor Late Infantile (left) and Juvenile onset (right). For each participant, the form(Vineland-IIor Vineland-3) with a longer follow-up period was selected, so that each participant appears only once per panel. Where Vineland-II scores were used, the markers are black, and the markers are gray for Vineland-3. Solid lines connect observations from the same participant, but different participants are not otherwise demarcated. Dotted lines indicate normative expectations (population mean = 15, SD = 3).

**Figure S7c.**
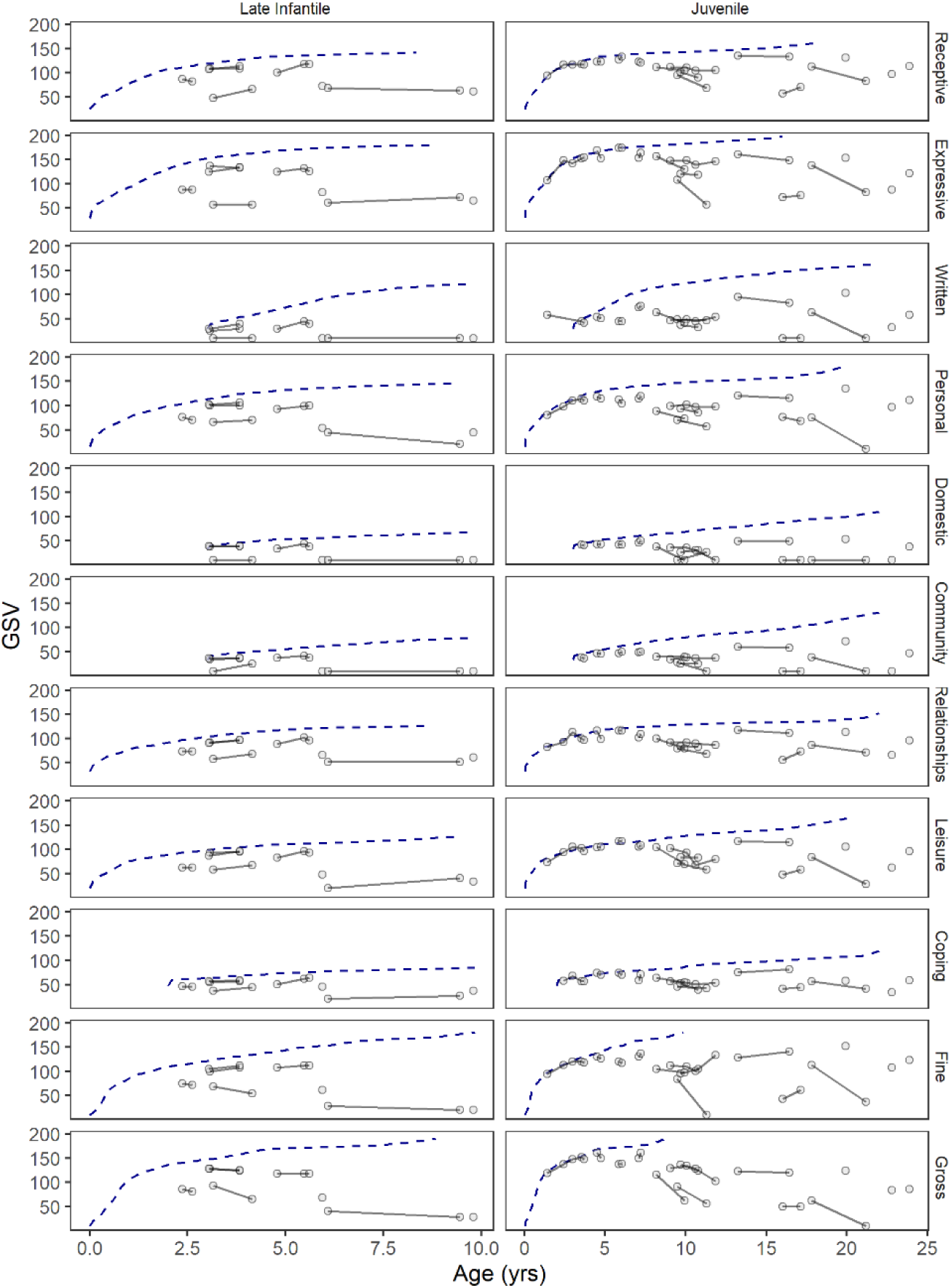
Longitudinal trends in Vineland subdomain-level growth scale values(GSVs) for Late Infantile (left) and Juvenile onset (right). Unlike the preceding plots 3a and 3b, all available Vineland-3 data are plotted. Solid lines connect observations from the same participant, but different participants are not otherwise demarcated. Dotted lines indicate normative expectations (median value per age equivalent).

**Figure S8.**
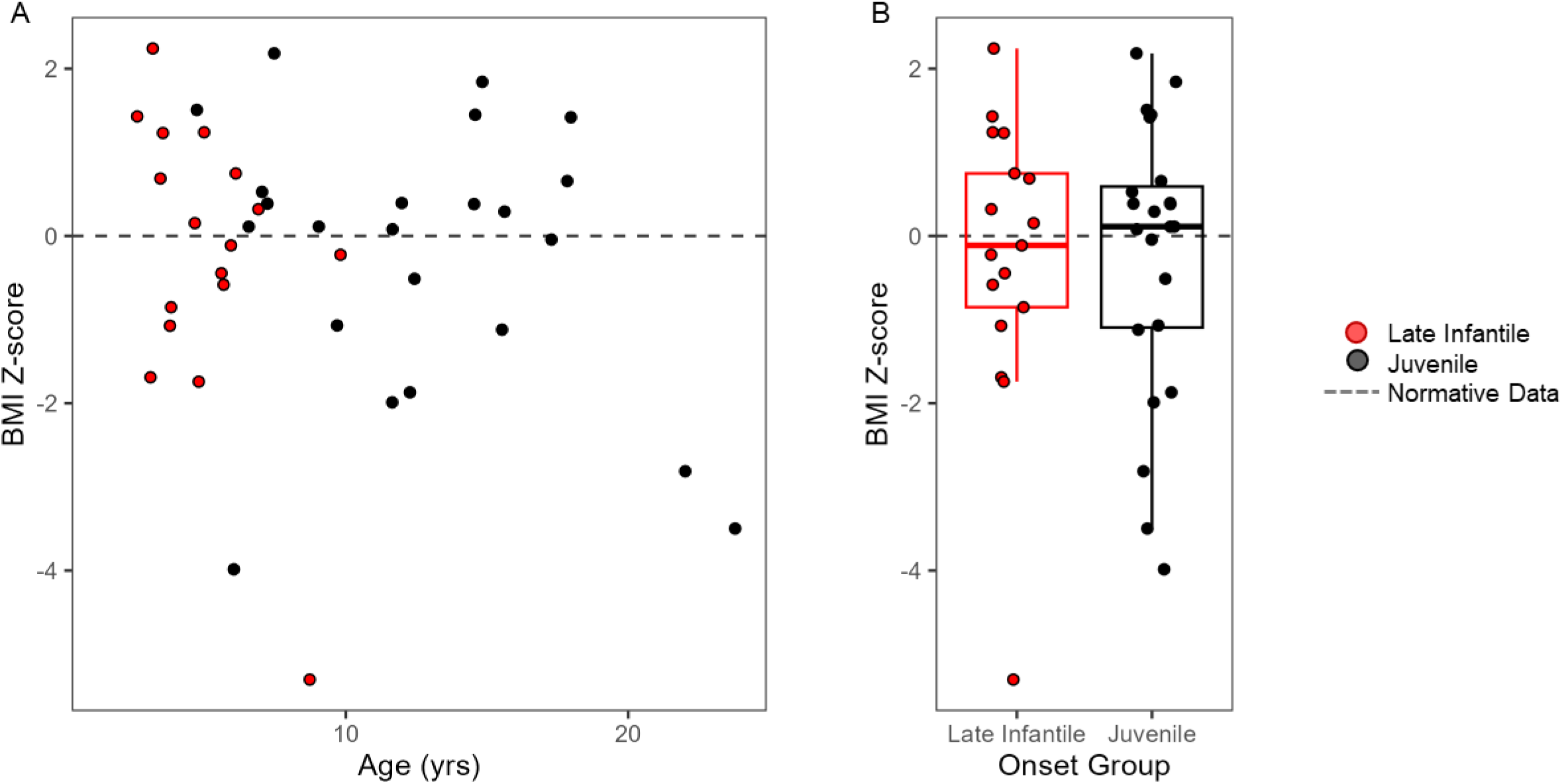
Age- and sex-based BMI Z-scores at the first assessment. Panel A: BMI Z-scores plotted by age. Panel B: BMI Z-scores plotted by group. Both panels: Red = late infantile onset, black = juvenile onset. CDC normative data for age 2 – 20 years were used; Z-score was not calculated for one participant aged 1.5 years and for two participants aged >20 years the 20-year norms were used.

